# Meditation, Psychedelics, and Brain Connectivity: A Randomised Controlled Resting-State fMRI Study of *N,N*-Dimethyltryptamine and Harmine in a Meditation Retreat

**DOI:** 10.1101/2025.07.30.25332422

**Authors:** Klemens Egger, Daniel Meling, Firuze Polat, Erich Seifritz, Mihai Avram, Milan Scheidegger

## Abstract

Both meditation and psychedelics are widely studied for their therapeutic potential in mental health. Recent research suggests potential synergies between mindfulness practice and psychedelics, though empirical studies have primarily focused on psilocybin. This study investigates the distinct and combined effects of mindfulness practice and an ayahuasca-inspired formulation containing *N,N*-dimethyltryptamine (DMT) and harmine on brain functional connectivity (FC), with implications for advancing clinical interventions. In this double-blind, placebo-controlled pharmaco-fMRI study, 40 meditation practitioners participated in a three-day meditation retreat. They were randomized to receive either placebo or buccal DMT-harmine (120 mg each) and underwent fMRI scans two days before and after administration. Neural changes were assessed using multiple connectivity metrics, including within- and between-network connectivity, network and global connectivity, and cortical gradient analyses. Within-group changes showed that meditators in the placebo group exhibited increased network segregation across several resting-state networks, while the DMT-harmine group showed increased FC within the visual network (VIS) and between VIS and attention networks. Between-group differences similarly showed increased FC between VIS and the salience network (SAL) in the DMT-harmine group compared to placebo post-retreat. No evidence of prolonged cortical gradient disruption, which is characteristic of acute psychedelic action, was observed. This suggests a return to typical brain organization shortly after the experience. These findings reveal distinct neural mechanisms underlying meditation and psychedelic-augmented meditation. While meditation alone reduced FC between networks, DMT-harmine increased within- and between-network connectivity. Given the potential of meditation and psychedelics for improving mental health, further exploration of their synergistic potential in clinical contexts is warranted. This study advances the understanding of how psychedelics and mindfulness practice shape brain function, offering insights into their complementary roles in emotional and psychological well-being.

## 1. Introduction

Serotonergic psychedelics, such as psilocybin, mescaline, LSD, and N,N-dimethyltryptamine (DMT), are compounds that can profoundly alter consciousness by inducing changes in perception, mood, cognition, and self-awareness that are often accompanied by mystical-type or peak experiences. These effects are induced mainly via agonism at the serotonin 2A receptor (5-HT_2A_R) (Nichols, 2016; Vollenweider & Preller, 2020). Ayahuasca, a traditional plant-based brew used by Indigenous Amazonian cultures for ritualistic purposes, combines DMT with the monoamine oxidase type A inhibitor (MAOI) harmine, enabling oral activity of DMT and producing intense visionary experiences (Dominguez-Clave et al., 2016; Riba et al., 2012). Ayahuasca gained attention for its therapeutic potential in treating mental health conditions like depression, post-traumatic stress disorder (PTSD), and addiction, as part of a broader resurgence of interest in psychedelics for therapy (Dominguez-Clave et al., 2016; Maia et al., 2023; Thomas et al., 2013; Uthaug et al., 2021). This therapeutic potential has recently inspired the development of novel ayahuasca-like formulations (Dornbierer et al., 2023; Egger et al., 2024). Furthermore, recent research suggests that the psychedelic effects of ayahuasca are primarily driven by DMT plasma concentrations, and short-acting DMT formulations—such as those administered via inhalation—are currently being investigated for their potential antidepressant effects (Falchi-Carvalho et al., 2025; Ramaekers et al., 2023, 2025).

On the other hand, various meditation practices have also been shown to facilitate altered states and traits of consciousness and have gained significant research attention in past years (Davidson, 2021; Van Dam et al., 2018). Defined as mental training aimed at promoting human flourishing, meditation encompasses a variety of practices that target core dimensions of well-being, including awareness, connection, insight, and purpose (Dahl et al., 2020). Both psychedelics and meditation have demonstrated positive effects on mental health (Andersen et al., 2021; Goldberg et al., 2018; Muttoni et al., 2019; Romeo et al., 2020; Rose et al., 2020), and recent efforts have sought to explore their overlaps, differences, and possible synergistic effects (Griffiths et al., 2018; Meling et al., 2024; Milliere et al., 2018; Moujaes et al., 2024; Smigielski et al., 2019).

To better understand the neural basis underlying the altered states of consciousness induced by meditation or psychedelics, functional magnetic resonance imaging (fMRI) has been used to track changes in functional connectivity (FC) patterns during and after such experiences. Studies of *acute* psychedelic states have repeatedly shown altered patterns of brain connectivity, characterized by increased desynchronization, signal diversity or entropy (Carhart-Harris et al., 2014; Lebedev et al., 2016; Siegel et al., 2024), reduced regional modularity (i.e., greater between-network FC) and lower within-network FC in certain networks (e.g., visual network (VIS) and default mode network (DMN)) (Avram et al., 2024; Mallaroni et al., 2024; Siegel et al., 2024; Timmermann et al., 2023). Some fMRI studies also investigated *subacute* effects of ayahuasca, typically 24 h after administration. These studies reported increased integrity in the salience network (SAL) but decreased integrity in DMN, while between-network FC was increased between SAL-DMN and between VIS-DMN but reduced between VIS-SAL (Pasquini et al., 2020; Sampedro et al., 2017). Somewhat distinct patterns have been observed in research on meditation—particularly mindfulness— with reports of reduced connectivity between VIS-DMN, which is thought to enhance self-awareness during and after mindfulness training (Moujaes et al., 2024; Sezer et al., 2022). Meditation has also been shown to increase connectivity between DMN and the frontoparietal network (FPN), possibly explaining improved attentional control in meditators (Sezer et al., 2022). A recent meditation retreat study found increased SAL-DMN FC and increased SAL integrity after an 8-day retreat in meditators (Vishnubhotla et al., 2021).

Furthermore, two studies explored the combined effects of psychedelics and meditation using fMRI, by examining the post-acute effects of psilocybin versus placebo administered during meditation retreats within the same study cohort (Singer et al., 2024; Smigielski et al., 2019). One of these studies investigated the effects of meditation and psychedelic-augmented meditation on FC metrics. Specifically, Smigielski and colleagues demonstrated both increased and decreased within-DMN connectivity in distinct DMN regions following psilocybin: increased FC in the frontal DMN (i.e., medial prefrontal cortex (mPFC) and anterior cingulate cortex), and reduced FC between mPFC and posterior cingulate cortex (PCC) and angular gyrus (Smigielski et al., 2019). While these findings suggest potential synergistic effects between psychedelics and meditation, the neural correlates and condition-specific differences have not been systematically investigated, and it is unclear if these changes are unique to psilocybin. To extend this line of research, we conducted a study with meditation practitioners in a Zen retreat center in the Swiss mountains to examine the combined effects of DMT and harmine with meditation over a three-day retreat. First results from psychometric assessments of the retreat have been published recently (Meling et al., 2024). Herein, we focus on changes induced by meditation and psychedelic-augmented meditation in brain network connectivity.

Specifically, we examined changes in resting-state FC both within groups (post vs pre retreat fMRI scans) and between groups (DMT-harmine vs placebo). Analyses focused on four key areas: 1) within- and between-network FC of large-scale resting state networks (RSNs), 2) connectivity between these RSNs and the whole-brain, 3) voxel-wise global connectivity strength across the whole brain, and 4) cortical gradient hierarchy and dispersion, reflecting both the principal organization of information flow from unimodal sensorimotor cortices to transmodal integrative regions, and the variability or spread of functional embedding across this hierarchy (Bethlehem et al., 2020; Sydnor et al., 2021a). Based on prior research into acute and subacute effects of DMT and ayahuasca on brain connectivity (Mallaroni et al., 2024; Pasquini et al., 2020; Timmermann et al., 2023), we hypothesized that the DMT-harmine group would show increased within-network connectivity in SAL and lower within-network-connectivity in DMN, and increased between-network connectivity between VIS-DMN and SAL-DMN, compared to the placebo group and compared to before the retreat. We expected weaker or different effects in the placebo group such as decreased FC between VIS-DMN and increased FC between SAL-DMN, respectively (Moujaes et al., 2024; Sezer et al., 2022; Vishnubhotla et al., 2021). We also anticipated higher global FC in the DMT-harmine group. Given the evidence that various psychedelics like psilocybin, LSD, and DMT can alter cortical gradients (Girn et al., 2022; Timmermann et al., 2023), we assessed whether changes in cortical information processing hierarchy persisted two days after DMT-harmine administration.

## 2. Subjects and Methods

This study was conducted in accordance with the Declaration of Helsinki and International Conference on Harmonization Guidelines in Good Clinical Practice and was approved by the Cantonal Ethics Committee of the Canton of Zürich (BASEC-Nr. 2021-00180). It also received an exemption from the Federal Office of Public Health (FOPH) for the administration of the controlled substance DMT. The study was registered at ClinicalTriails.gov (<u>NCT05780216</u>). All participants provided written informed consent.

### 2.1. Participants and experimental procedures

This study was conducted using a double-blind, placebo-controlled, between-within subject mixed design (**Fig. 1**). 40 healthy meditation practitioners participated in one of two structurally identical three-day meditation retreats (n=19 and n=21, respectively). Participants were screened in advance and, upon successful enrollment, randomly allocated to receive either placebo or received either placebo or DMT-harmine on the second retreat day (details on the study drug formulation are given below).

**Figure 1.**
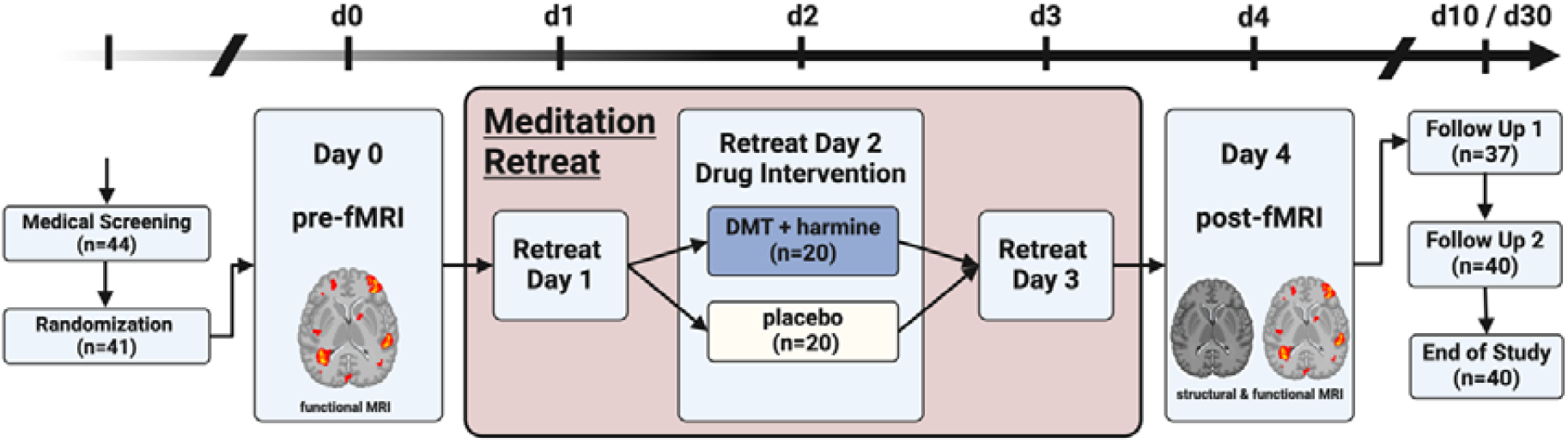
Study design. Double-blind, placebo-controlled, mixed design with pre- and post-retreat MRI assessments. Forty-one meditation practitioners were randomized to receive either DMT-harmine or placebo during a 3-day meditation retreat; 40 participated in and completed the study. Psychometric assessments were conducted throughout the retreat and at follow-up. Refer to text and the original publication for details (Meling et al., 2024).

To accommodate scanner availability, each retreat was divided into two cohorts that started one day apart, allowing for MRI scanning of up to 11 participants per day. The first cohort began on Day 0 and ended on Day 4; the second cohort began on Day 1 and finished on Day 5. One day before the retreat, all participants underwent fMRI scanning at the University Hospital of Psychiatry Zurich (“Pre” assessment). On the second retreat day, the pharmacological intervention took place. On the day following the retreat (“Post” assessment), participants returned to the hospital for a follow-up MRI session, including structural and functional sequences.

Throughout the retreat, participants engaged in full days of meditation practice (e.g., sitting and walking meditation). Subjective experiences were assessed each evening using the Mystical Experience Questionnaire (MEQ) (Barrett et al., 2015). Additional questionnaires included the Nondual Awareness Dimensional Assessment-State (NADA-S), the Toronto Mindfulness Scale (TMS) (Lau et al., 2006), the Sussex-Oxford Compassion Scales for Self and Other (SOCS-S and SOCS-O) (Gu et al., 2020), the Psychological Insight Scale (PIS) (Peill et al., 2022) and the Emotional Breakthrough Inventory (EBI) (Roseman et al., 2019) to cover the four dimensions 1) mindfulness, 2) compassion, 3) insight, and 4) transcendence that were investigated in the original study. One week and one month after the retreat, participants additionally filled out psychometric questionnaires. For further information on psychometric assessments and retreat structure, refer to the original publication (Meling et al., 2024).

Out of the 40 participants, data from 39 participants were analyzed herein (22 male, 17 female; 19 DMT-harmine, 20 placebo, mean age 43.5 ± 10.3 years; mean meditation experience 2450 ± 1890 hours, refer to **Suppl. Table 1** for details). One participant was excluded from the analyses due to excessive head motion during functional scans (refer to **Suppl. Table 2**). Aside from one participant who reported headaches after DMT+harmine administration, no major adverse events occurred during the study.

### 2.2. Substance and dosing

The pharmacological intervention utilized a standardized oromucosal formulation containing DMT and harmine as described elsewhere (Egger et al., 2025; Meling et al., 2024). The formulation included calcium phosphate template-inverted particles (TIP) (Kost et al., 2023), which were separately loaded with either 25% DMT or 25% harmine (freebase weight). These DMT- and harmine-loaded TIP particles were then combined into the same tablets, yielding 1:1 ratio (freebase weight) of DMT and harmine per tablet. Each tablet also contained 3% croscarmellose sodium, along with 0.5% sucralose, 0.5% menthol, and 0.5% peppermint flavor for taste masking. Each tablet contained 30⍰mg of DMT and 30⍰mg of harmine (both as freebase weight), with a total of four tablets administered at 30-minute intervals (Meling et al., 2024). This resulted in a cumulative dose of 120 mg DMT and 120 mg harmine. Placebo tablets were manufactured using the same formulation process, but containing TIP particles that were not loaded with DMT or harmine. All other ingredients were identical to those in the active tablets with the addition of a small amount of Bitrex^®^ (denatonium benzoate) to ensure matching appearance and taste. Despite this, >80% of the participants correctly identified their drug allocation at the end of the dosing day (Meling et al., 2024), thus limiting blinding efficacy (Aday et al., 2025).

### 2.3. Data acquisition and preprocessing

Structural and functional MR images were acquired on a 3T MR scanner (Achieva 3.0T, Philips, Amsterdam, The Netherlands) equipped with a 32-channel receive head coil and MultiTransmit parallel radio frequency transmission was used. 10–11 participants were scanned per scanning day (two pre- and post-scanning days per retreat). A T1-weighted image was obtained only on the second imaging visit with a 3D multishot Turbo Field Echo (TFE) sequence with the following specifications: repetition time (TR)=8.2 ms, echo time (TE)=3.8 ms, flip angle (FA)=8°, field-of-view (FoV)=240×240 mm^2^, slices=160, no interslice gap, voxel size=1.0×1.0×1.0 mm^3^, acquisition time (TA)=4.53 min. Functional images were acquired on both pre- and post-retreat sessions with a resting-state protocol (eyes closed) using a whole-brain gradient echo planar imaging (EPI) sequence with the following specifications: TR=1800 ms, TE=35 ms, FA=70°, FoV=220×220 mm^2^, matrix size=124×121 mm^2^, slices=54, no interslice gap, voxel size=1.7×1.7×2.0 mm^3^, number of scanned volumes=240, TA=7.40 min.

Preprocessing was performed using the configurable *fMRIPrep* 23.0.2 (Esteban et al., 2019, 2023) pipeline, which included intensity non-uniformity correction, skull-stripping, and spatial normalization to *MNI152NLin2009cAsym* space. Functional data preprocessing steps included motion correction, slice-timing correction, and co-registration to the T1-weighted image. Further, denoising steps were performed with *CONN* (Whitfield-Gabrieli & Nieto-Castanon, 2012) release 22.a (Nieto-Castanon & Whitfield-Gabrieli, 2022) and included smoothing of the data with a 6 mm FWMH Gaussian kernel and regression of motion artifacts with the Friston 24-parameter model (Friston et al., 1996), physiological noise correction with the aCompCor50 method including combined white matter and cerebrospinal fluid components explaining 50% of the variance per scan (Behzadi et al., 2007), scrubbing of volumes with a framewise displacement (FD) > 0.5 mm (Power et al., 2014), and temporal filtering (0.008–0.09 Hz) (Hallquist et al., 2013). Additionally, the regression of global signal (GSR) was done as complementary analysis, and results are reported both with (in Supplement) and without GSR as the inclusion of GSR is still an ongoing debate in the (psychedelic) neuroimaging field (Avram et al., 2024; McCulloch et al., 2022). For some analyses (as described below), fully preprocessed and denoised data were transformed from *MNI152NLin2009cAsym* volumetric space into *fsaverage5* surface space using *surface*.*vol_to_surf* (Nilearn 0.10.4). A detailed description of each step is provided in the Supplementary Methods.

### 2.4. Network connectivity

The RSNs used herein were based on the templates of Yeo and colleagues and included the visual network (VIS), auditory-sensorimotor network (ASM), dorsal attention network (DAN), salience network (SAL), frontoparietal network (FPN), and the default-mode network (DMN) (visualized in **Fig. 3, Panel A**) (Yeo et al., 2011). We excluded the limbic network (LIM) from our analyses given its comparably lower reproducibility and signal-to-noise ratio (Avram et al., 2024; Gordon et al., 2016; Yeo et al., 2014). Network FC maps were computed with *CONN*. Seed regions included the above-mentioned RSNs (Yeo et al., 2011). Network connectivity was conducted to calculate FC between each RSN and every gray matter voxel in the brain. FC reflects Fisher-transformed bivariate correlation coefficients from a weighted general linear model (weighted-GLM (Nieto-Castanon, 2020)), estimated separately for each seed area (i.e., one of the 6 RSNs) and target voxel, modeling the association between their BOLD signal timeseries.

### 2.5. Within- and between-network connectivity

Within-network FC was calculated using first-level maps obtained from the network connectivity analysis described above. For each RSN, these subject-specific connectivity maps were masked with the corresponding network mask to extract the average FC within that network using *fslmeants*. This yielded subject- and condition-specific FC values for each RSN.

RSN-to-RSN FC (between-network FC) matrices were estimated characterizing FC between each pair of regions among the 6 RSNs with *CONN’s* ROI-to-ROI FC analysis option. FC strength was estimated separately for each pair of ROIs (i.e., RSNs), characterizing the association between their BOLD signal timeseries.

### 2.6. Global Connectivity analysis

Global FC was computed using the *BrainSpace* toolbox (0.1.10, Python implementation (Vos de Wael et al., 2020)), with data analysis performed in *fsaverage5* surface space. To reduce data dimensionality, data in *fsaverage5* space (20,484 vertices) was parcellated into 400 regions using the Schaefer atlas (Schaefer et al., 2018), by averaging the values of all vertices within each parcellation. For each scan, a 400×400 functional connectivity matrix was computed by calculating Pearson’s correlation coefficients between the time series of all pairs of Schaefer parcels. These correlation coefficients were then Fisher z-transformed. Global FC was defined as the average of each parcel’s correlation with all other parcels, resulting in one global connectivity value per region and scan. Similar calculations of global connectivity were performed with other psychedelic substances such as LSD, psilocybin, and DMT during acute states (Madsen et al., 2021; Preller et al., 2018, 2020; Timmermann et al., 2023).

### 2.7. Cortical gradient analysis

Cortical gradient analysis used the same Fisher z-transformed FC matrices calculated for global FC as a starting point. These individual connectivity matrices were subjected to row-wise thresholding to only maintain the top 10% of edges per row, as has been done previously (Bethlehem et al., 2020; Girn et al., 2022; Timmermann et al., 2023). Subsequently, cosine similarity was calculated to generate a similarity matrix that served as input for the diffusion map embedding algorithm, a nonlinear manifold learning method that uses graph Laplacians to identify gradient components at the individual subject level (Bethlehem et al., 2020; Coifman et al., 2005; Margulies et al., 2016). Diffusion map embedding reduces the high-dimensional similarity matrix into a low-dimensional space of embedding components (gradients).

Each gradient represents a dimension of FC pattern similarity, where vertices with many or strong connections are positioned closer together in the embedding space, and vertices with weaker or fewer connections are farther apart. The Euclidean distance in this space reflects the “difference in gradient scores” between regions. A single parameter α controls the influence of data point density on the embedding manifold (α = 0 indicates maximum influence; α = 1, no influence). Diffusion map embedding is characterized by α = 0.5 (Coifman et al., 2005), which balances the consideration of both global and local data point relationships in determining the embedding space.

To enable comparisons between subjects, Procrustes rotation was applied after gradient estimation to align individual gradient components to an all-subjects-baseline group average embedding template with 10 iterations (Bethlehem et al., 2020; Girn et al., 2022; Vos de Wael et al., 2020). This alignment ensures that gradient axes are consistent across scans and individuals. Heat maps showing Pearson correlations of individual scans of the first three gradients with the embedding template after Procrustes rotation can be found in **Suppl. Figure 3**. We calculated the first 10 cortical gradients with this approach, but only used the first three gradients for further analysis, as those already explained ∼70% of the variance in the data (DMT-harmine pre scans: 69.4%, placebo pre scans: 71.2%, DMT-harmine post scans: 70.5%, and placebo post scans: 69.6%). These three main gradients were visualized and compared between groups and timepoints. To further characterize multi-dimensional differences in hierarchical organization in cortex, we calculated within- and between-network “dispersion” in 3D space (Bethlehem et al., 2020). Within-network dispersion was calculated as the sum squared Euclidean distance of network nodes (parcellations) to the centroid of this network at the individual scan level. Between-network dispersion represents the Euclidean distance between network centroids (Bethlehem et al., 2020).

### 2.8. Statistical Analyses

The following four within- and between-group contrasts were defined and used as second level contrasts for each connectivity measure: (i) **Synergy** (DMT-harmine > Placebo: Post > Pre), representing the group-by-time interaction that reflects the added effect of the drug to the intervention vis-à-vis group differences from baseline, i.e., synergistic effects; (ii) **Group differences post retreat** (DMT-harmine > Placebo: Post), that evaluates the effect of DMT-harmine against placebo, including drug+meditation interactions; (iii) Meditation (Placebo: Post > Pre) reflecting the effect of the meditation retreat under placebo; and (iv) **Psychedelic-augmented meditation** (DMT-harmine: Post > Pre), reflecting combined effect of DMT-harmine and meditation.

For within- and between-network FC analyses, the first contrast was implemented as a two-way repeated measures ANOVA with drug allocation (DMT-harmine > placebo) as between-subject and time (post > pre) as within-subject factor, the second as a between-group ANCOVA comparison of post-intervention scores, with pre-intervention values included as covariates, and the last two as ANCOVAs restricted to the corresponding group, with baseline values of the other group included as covariates.

Network connectivity group level analyses were performed using a General Linear Model (GLM (Nieto-Castanon, 2020)). For each individual voxel a separate GLM was estimated, with first-level connectivity measures at this voxel as dependent variables, and group and/or session as independent variables. Voxel-level hypotheses were evaluated using multivariate parametric statistics with random-effects across subjects and sample covariance estimation across multiple measurements. Inferences were performed at the level of individual clusters. Cluster-level inferences were based on parametric statistics from Gaussian Random Field theory (Nieto-Castanon, 2020; Worsley et al., 1996).

Group contrasts for global FC and gradient analysis, including gradient dispersion, were conducted with the *SLM* function for surface-based linear models (*BrainStat* 0.4.2, (Larivière et al., 2023)).

If not stated otherwise, all results for voxelwise analyses were thresholded using a combination of a cluster-forming *p*<.001 voxel-level threshold, and a FDR-adjusted *q*_FDR_<.05 cluster-size threshold (Chumbley et al., 2010). The corresponding smoothness estimates are reported in **Suppl. Table 5**.

We additionally evaluated associations between psychometric questionnaires that showed a significant between- or within-group effect at Day 2 (i.e., MEQ, EBI, NADA-S, PIS, and TMS) as reported in (Meling et al., 2024) and the average timeseries of the significant cluster from the **Group differences post retreat** contrast that resulted from the network connectivity analysis using ordinary least squares regression. This cluster was chosen as it denotes consistent and specific FC changes in comparison to the placebo group (see below). The average timeseries derived from the cluster was used as the dependent variable and the questionnaire scores and group (i.e., drug allocation) were used as predictors, including main effects and interaction. We used FDR-correction to account for the number of questionnaires modeled. Significant interactions were visualized with a grouped scatter plot with the corresponding Person’s / Spearman rank correlations.

To further explore the relationship between changes in subjective experience and FC, we repeated the association analysis using delta scores for each questionnaire to control for individual baseline differences, calculated as the difference between Day 2 (post-drug intervention) and the respective baseline values (assessed on Day 1, i.e. in the evening of the first retreat day).

## 3. Results

### 3.1. Network connectivity-based correlation analysis

Voxel-wise FC differences between groups and timepoints were investigated for each RSN with the whole brain (**Fig. 2**).

**Figure 2.**
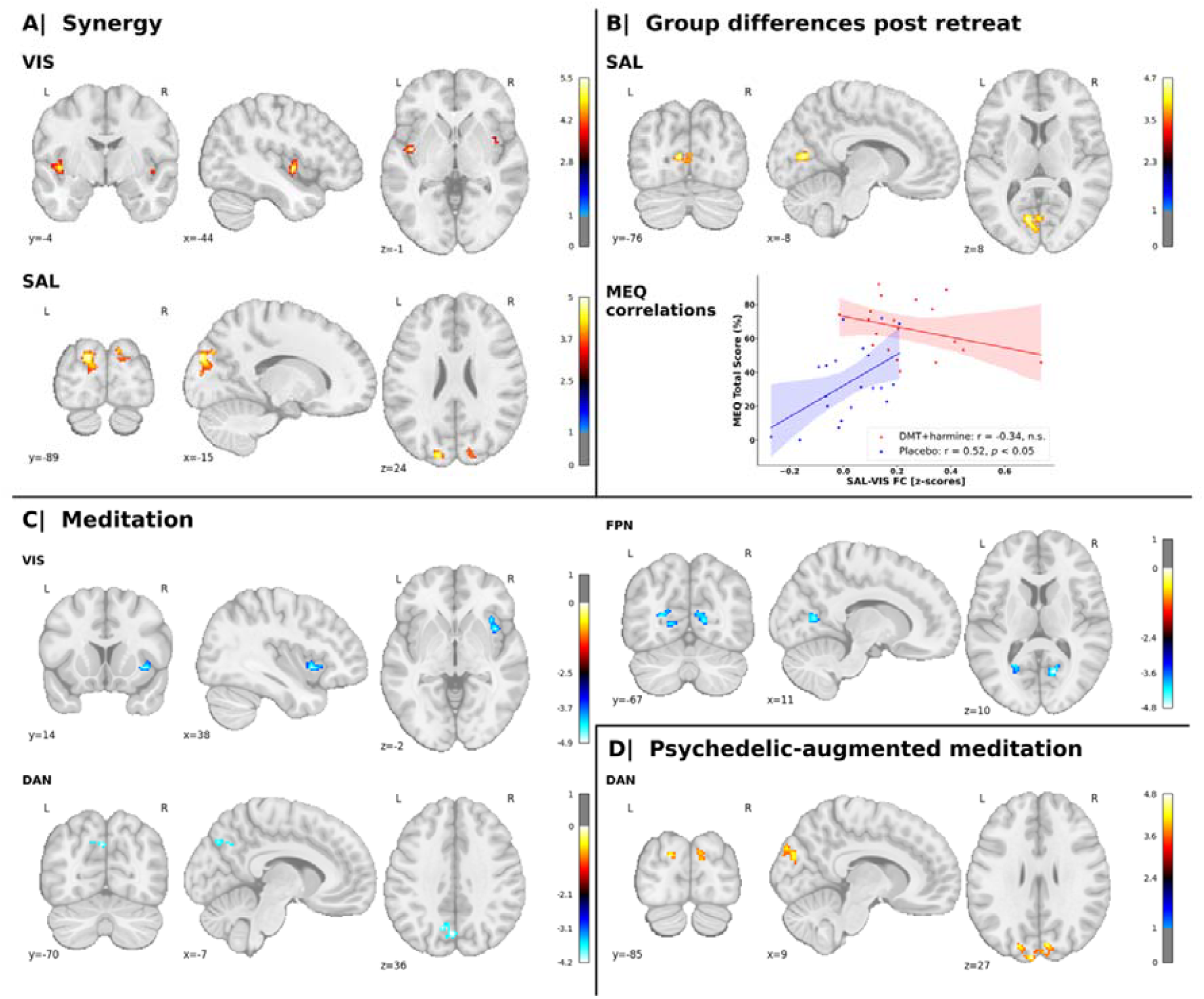
Changes in network connectivity between groups and timepoints. Voxel-wise statistical parametric maps that represent changes in connectivity between one RSN (i.e., the seed) and the whole brain are shown. The allocation of each significant cluster to its corresponding Yeo network is indicated in parentheses. The panels correspond to different contrasts between drug condition (i.e., DMT-harmine vs placebo) and timepoint (i.e., pre vs post meditation retreat). **A**| **Synergy** (DMT-harmine > placebo: Post > pre): The two-way repeated measures ANOVA model shows increased FC between VIS and bilateral insula (SAL) and between SAL and bilateral occipital poles and cuneus (VIS) for the DMT-harmine group. **B**| **Group differences post retreat** (DMT-harmine > Placebo: Post): After the retreat, an ANCOVA showed increased FC between SAL and bilateral calcarine sulcus (VIS) in the DMT-harmine group compared to placebo. This cluster showed significant group interactions with the Mystical Experience Questionnaire (MEQ) on Day 2. Pearson’s or Spearman’s rank correlations between the average connectivity change in this cluster and MEQ scores are shown. MEQ scores were significantly correlated with SAL-VIS FC scores for the placebo (blue), but not for the DMT-harmine group (red). **C**| **Meditation** (Placebo: Post > Pre): ANCOVA indicated FC decreases within the placebo group after the retreat between VIS and right insula (SAL), between DAN and precuneus (DMN), and between FPN and bilateral cuneus, lingual gyri, and calcarine cortex (VIS). **D**| **Psychedelic-augmented meditation** (DMT-harmine: Post > Pre): ANCOVA demonstrated increased FC between DAN and bilateral occipital pole (VIS). The analyses were computed in CONN (*p*⍰< ⍰.001, cluster-level FDR-adjusted *q*_FDR_⍰< ⍰.05); x, y, and z indicate Montreal Neurological Institute (MNI) coordinates; warm colors represent increased connectivity and cold colors decreased connectivity for the given contrast. Refer to **Suppl. Fig**. 1 for results with global signal regression.

i. With the **Synergy** contrast (*DMT-harmine* > *Placebo: Post > Pre*), we observed increased FC between VIS and bilateral insula (SAL) and between SAL and bilateral cuneus and occipital poles (VIS).
ii. **Group differences post retreat** (*DMT-harmine > Placebo: Post*) showed increased FC between the SAL and bilateral calcarine sulcus, part of the VIS.
iii. In the **Meditation** contrast (*Placebo: Post > Pre*), VIS showed decreased FC with the right insula of the SAL. Additionally, we found decreased FC between DAN and precuneus (part of DMN). Finally, the FPN displayed decreased FC with visual areas such as the bilateral cuneus, lingual gyri and calcarine cortex.
iv. In the **Psychedelic-augmented meditation** contrast (DMT-harmine: *Post > Pre*) we observed increased FC between DAN and bilateral occipital poles—reflecting increased DAN-VIS FC.

Results from complementary analyses conducted with GSR broadly overlap with these results, refer to **Suppl. Fig. 1** for details.

### 3.2. Within- and between network connectivity

For within- and between network connectivity, no significant changes were observed following correction for multiple comparisons for any contrast of interest (both with and without inclusion of GSR). However, several trends toward significance were observed without FDR correction.

In detail, for within-network FC, the **Synergy** contrast did not show significant interaction effects (*q*_FDR_<.05), but revealed a main effect of time in VIS (*F*(1,74)=6.16, *p*_uncorrected_=.015). Analysis of the other contrasts revealed a significant increase in within-network FC in VIS following the retreat in the DMT-harmine group (*t*=2.29, *p*_uncorrected_=.025) in the **Psychedelic-augmented meditation** contrast (**Fig. 3, Panel B**). No other RSN showed changed within-network FC. By including GSR as a denoising procedure, we observed a significant effect of time in VIS (*F*(1,74)=7.77, *p*_uncorrected_=.007) for the Synergy contrast. The **Meditation** contrast showed increased within-network FC after the retreat (*t*=2.20, *p*_uncorrected_=.031) in VIS. No other contrast/network showed significant changes, see **Suppl. Table 6** and **Suppl. Fig. 2** for details.

**Figure 3.**
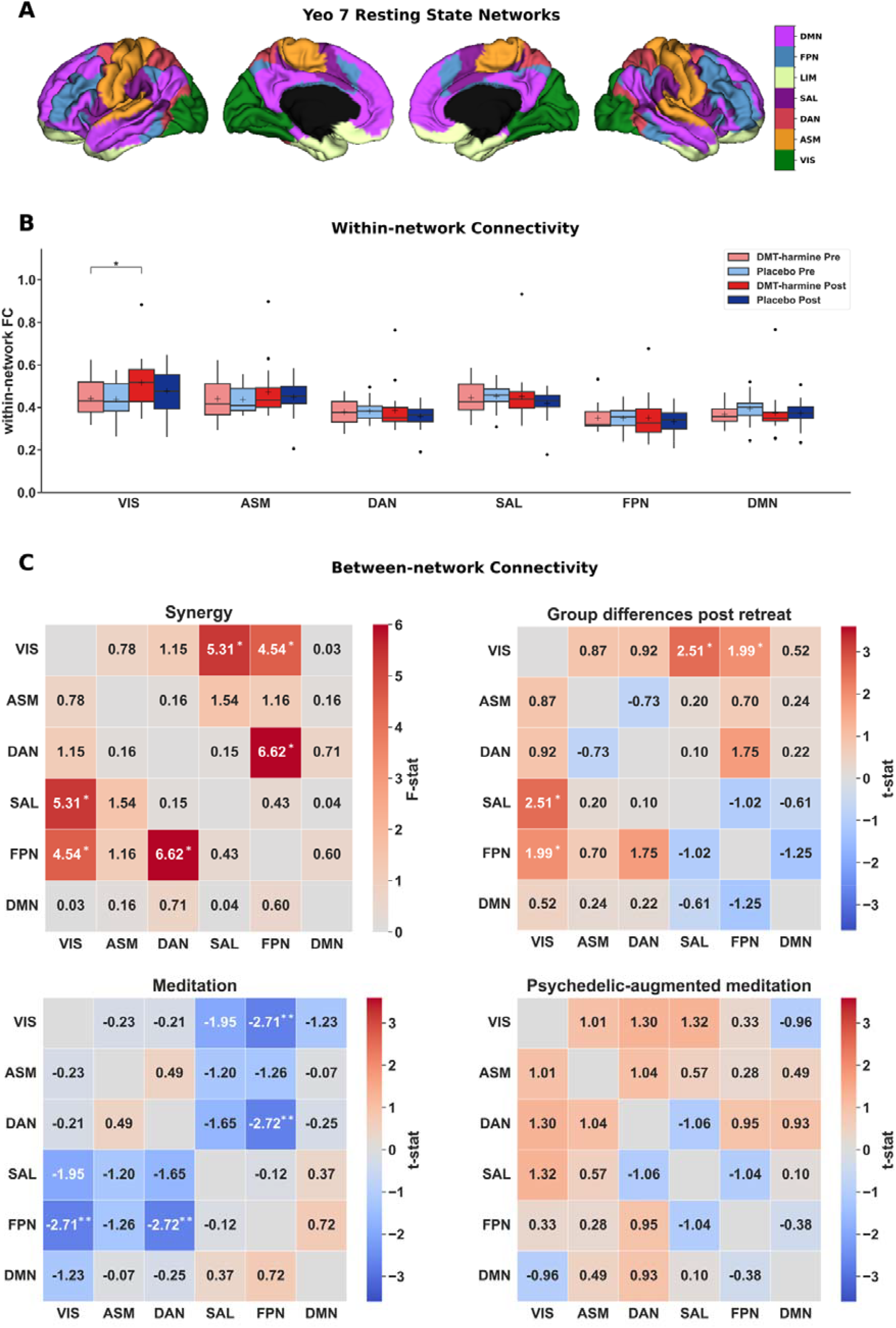
Within-and between-network functional connectivity (FC) changes after DMT-harmine or placebo. **A**| The seven canonical resting state networks defined by Yeo and colleagues plotted on cortical surfaces (Yeo et al., 2011). First two sections represent the left hemisphere, the last two sections represent the right hemisphere. The limbic network (LIM) was not used for analyses. **B**| No significant results were observed for the within-network FC analysis following multiple correction; however, the following trends were observed at the uncorrected level. Within-network FC analysis showed a significant main effect of time in VIS in the Synergy contrast (*F*(1,74)=6.16, *p*_uncorrected_=.015). A significant increase in within-VIS FC was observed following the retreat in the DMT-harmine group in the **Psychedelic-augmented meditation** contrast (*t*=2.29, *p*_uncorrected_ =.025). No other significant within-network effects were detected for the **Group differences post retreat** and **Meditation** contrasts. **C**| Similarly, for between-network FC analysis no results survived correction for multiple testing. However, uncorrected trend-level effects were observed. Repeated-measures ANOVA (**Synergy** contrast, top left matrix) showed significant interaction effects between VIS-SAL (*F*(1,74)=5.31, *p*_uncorrected_=.024), VIS-FPN (*F*(1,74)=4.54, *p*_uncorrected_=.037), and DAN-FPN (*F*(1,74)=6.62, *p*_uncorrected_=.012). In the **Group differences post retreat** contrast, increased FC was observed between VIS-SAL (*t*=2.51, *p*_uncorrected_=.014) and VIS-FPN (*t*=1.99, *p*_uncorrected_=.050) in the DMT-harmine group. The **Meditation** contrast showed decreased FC between VIS-FPN (*t*=2.71, *p*_uncorrected_=.001) and DAN-FPN (*t*=2.72, *p*_uncorrected_=.001) after the retreat. Results were consistent when including global signal regression (see **Suppl. Fig**. 2). Asterisks denote significant differences for differences in t-tests for the given contrast without adjustment for multiple comparisons (*: *p*_uncorrected_⍰<.05, **: *p*_uncorrected_ ⍰<.01). Abbreviations: VIS – visual network, ASM – auditory-sensorimotor network, DAN – dorsal attention network, SAL – salience network, FPN – frontoparietal network, DMN – default mode network.

Between-network FC analysis results are shown in **Fig. 3, Panel C**. The **Synergy** revealed interaction effects between VIS-SAL (*F*(1,74)=5.31, *p*_uncorrected_=.024), VIS-FPN (*F*(1,74)=4.54, *p*_uncorrected_=.037), and DAN-FPN (*F*(1,74)=6.62, *p*_uncorrected_=.012) connections. There was no significant effect in the **Psychedelic-augmented meditation** contrast. In the **Meditation** contrast, a decrease in FC between VIS-FPN (*t*=2.71, *p*_uncorrected_=.001) and DAN-FPN (*t*=2.72, *p*_uncorrected_=.001) was noted following the retreat. In the **Group Differences Post Retreat** contrast, increased FC was observed between VIS-SAL (*t*=2.51, *p*_uncorrected_=.014) and VIS-FPN (*t*=1.99, *p*_uncorrected_=.050) in the DMT-harmine group compared to placebo. Including GSR did not have a strong impact on the results, with the same effects observed at the uncorrected level (**Suppl. Fig. 2**).

### 3.3. Global connectivity analysis

We examined global FC differences across time and groups using the four predefined contrasts (Fig. 4), based on the 400-region Schaefer parcellation of the cortical surface. None of the contrasts Synergy, Group differences post retreat, or Psychedelic-augmented meditation showed statistically significant changes in global FC after FDR correction, regardless of whether GSR was applied. However, in the Meditation contrast, three clusters located in SAL /FPN networks showed significantly decreased global FC after the retreat in the analysis without GSR (FDR-corrected; Fig. 4). These effects were not observed when GSR was applied.

**Figure 4.**
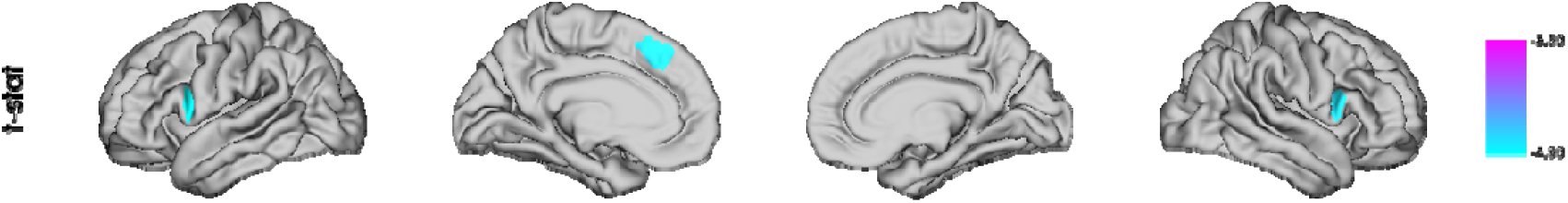
Global FC changes in the Meditation contrast. Results from an ANCOVA on global FC reveal three clusters with significantly decreased FC after the retreat in the **Meditation** contrast (*q*_FDR_<0.05). All significant clusters were located within the salience (SAL) and frontoparietal (FPN) networks.

### 3.4. Cortical Gradient Analysis

The cortical gradient analysis resulted in a hierarchical axis ranging from lower-level sensorimotor cortices (i.e., “lower” cognitive functions) to higher-level transmodal cortical regions (Gradient 1; **Fig. 5**), from insular to visual cortex (Gradient 2; **Suppl. Fig. 4**), and from visual cortex to cortices covered by the FPN regions (Gradient 3; **Suppl. Fig. 5**). We identified the typical cortical gradient axes previously reported (e.g., Bethlehem et al., 2020; Girn et al., 2022; Sydnor et al., 2021a) in all four scan conditions (*DMT-harmine Pre/Post, Placebo Pre/Post*). However, no significant differences were found between conditions or sessions in any brain region for any of the four defined contrasts. This was true both in surface space and when comparing network-wise gradient scores. Complementary analysis with inclusion of GSR resulted in the same gradient axes, but similar to the standard analysis, did not indicate any statistical differences between groups and conditions for any gradient (results not shown).

**Figure 5.**
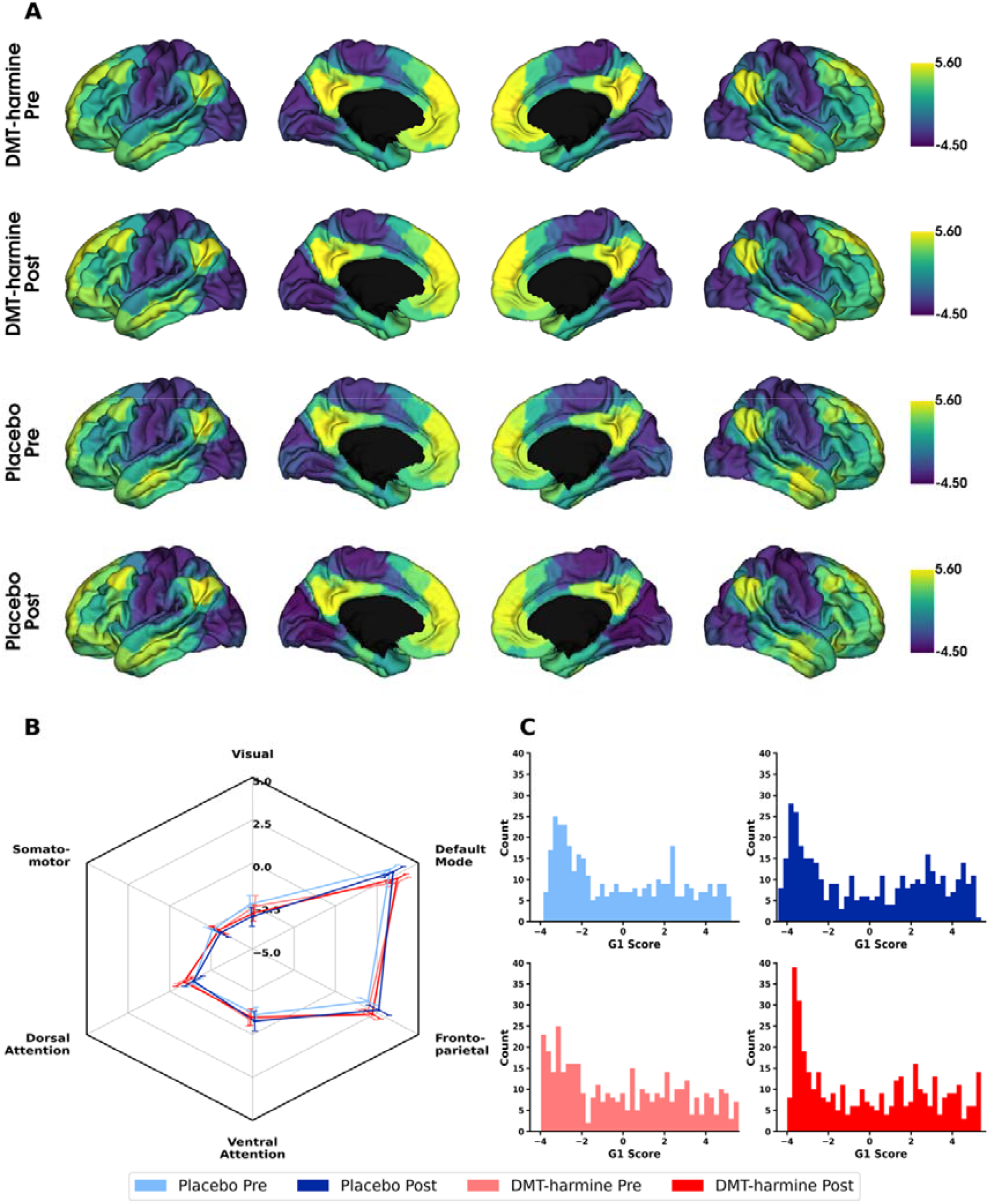
Cortical first gradients before and after retreat for DMT-harmine and placebo group. **A**| Parcellation-wise mean cortical principal gradients for DMT-harmine Pre, DMT-harmine Post, Placebo Pre and Placebo Post, conditions that represent the principal axis from unimodal (lower end) to transmodal (higher end) cortex. The shown gradient scores represent arbitrary units, with negative values defining the lower (sensorimotor) end of the gradient axis and higher (positive) numbers representing the higher (transmodal) end of the axis. **B**| Network-wise mean cortical gradients. The same data is presented network-based, wherein each brain parcel was allocated to the corresponding Yeo network. Mean and SEM are presented per network. **C**| Histograms showing the distribution of gradient values for each condition for each brain parcellation. Statistical differences were tested with a two-way repeated measures ANOVA (for within- and between group effects and their interactions). No significant differences were observed for any contrast, neither with parcellation-nor network-wise contrasts. Refer to **Suppl. Figs. 4 &** 5 for complementary figures showing the second and third cortical gradient.

Additional within- and between-network dispersion analyses in 3D gradient space did not indicate statistical differences between groups or timepoints (refer to **Suppl. Tables 7 & 8** for results from 2-by-2 ANOVA analysis of within- and between-network dispersion). Parcellation-wise gradient data is shown in **Fig. 6** in 3D and 2D gradient space (representing gradient values of each parcellation along all three gradients and each gradient pair combination). Similar to the gradient analysis, network dispersion analysis including GSR did not result in significant differences within- or between-networks (data not shown).

**Figure 6.**
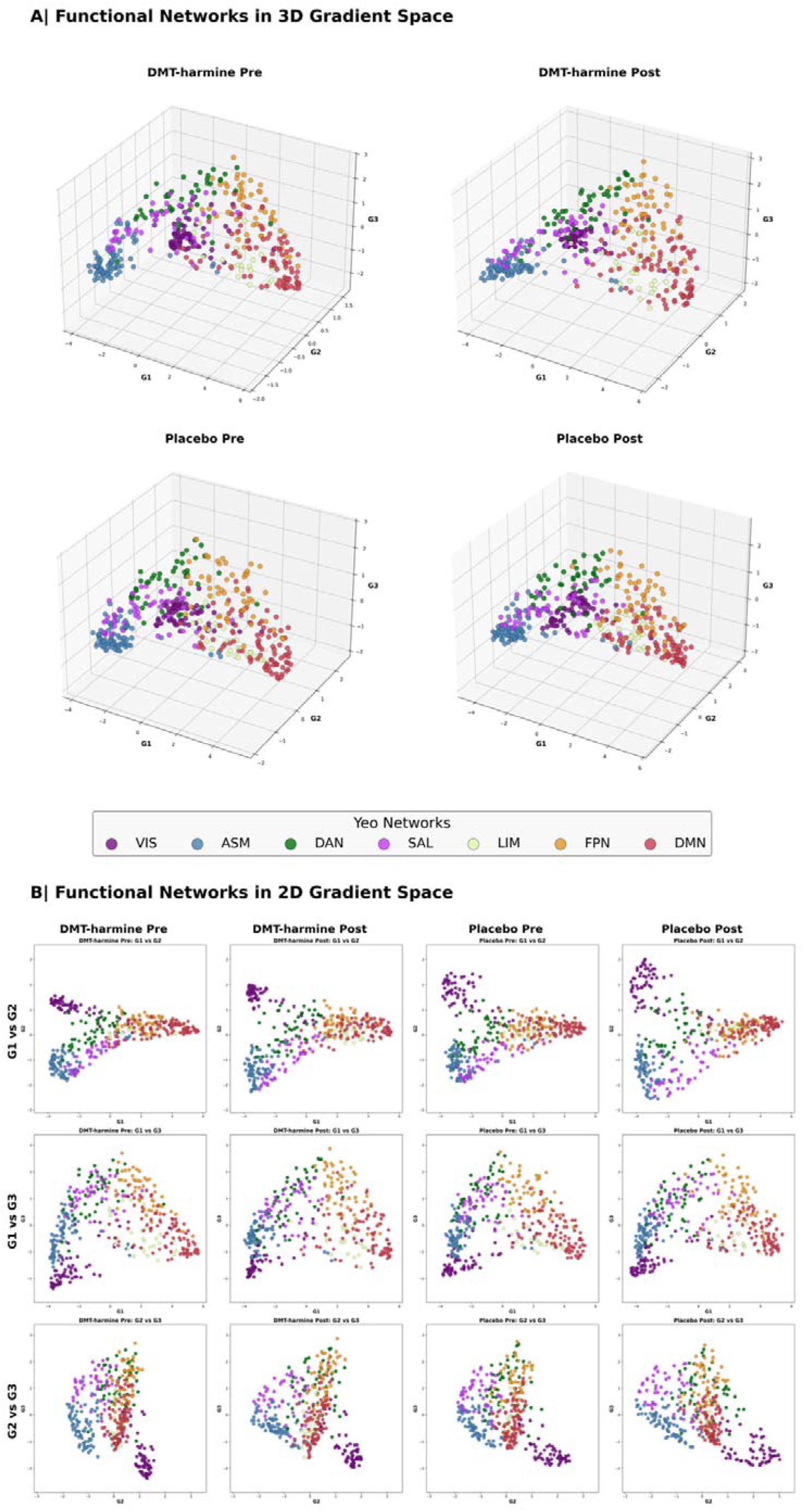
First three functional gradients in 3- and 2-dimensional gradient space. **A**| The first three functional gradients are projected into a three-dimensional coordinate system where each axis represents one of the cortical gradients. **B**| The same data presented in two-dimensions, with each combination of possible gradient pairs being displayed. Although there are slight differences in gradient mapping between groups observable by visual inspection, these differences did not result in statistical significance when comparing within- and between-network dispersion (refer to **Suppl. Tables 7 & 8** for ANOVA stats tables). Data presented in this figure is parcellated with the 400-region Schaefer Atlas, with each parcellation colored in the color of the Yeo network it belongs to.

### 3.5. Associations between network connectivity and psychometric outcomes

We used ordinary least squares regressions to explore the relationships between the cluster found for the **Group differences post retreat** contrast with the network connectivity analysis (increased connectivity between SAL- and bilateral calcarine sulci (i.e., VIS)) and questionnaires targeting mindfulness, emotional breakthrough, insight, and transcendence (Meling et al., 2024). We used FDR-correction for the total number of questionnaires used (n=5). Between-group differences were observed via significant interactions between SAL-VIS FC (contrast **Group differences post retreat; Fig. 2, Panel C**) and the questionnaires MEQ on Day 2 (*b*=0.0065, 95%CI [0.0014, 0.0117], *q*_FDR_=.036), and TMS on Day 2 (*b*=0.0089, 95%CI [0.0019, 0.0159], *q*_FDR_=.036). Post-hoc analyses revealed significant associations between SAL-VIS FC and MEQ on Day 2 for the placebo group (*r*=0.52, *p*=.020), while the TMS scores were not significantly correlated with SAL-VIS FC for any group (**Fig. 2**). To complement these findings, we repeated the association analysis using delta scores (Day 2 minus baseline) for each questionnaire. However, no significant group-by-questionnaire interactions were observed in the analysis of delta questionnaire scores. Refer to **Suppl. Tables 9–12** for OLS model and exploratory correlation results for all tested questionnaires.

## 4. Discussion

In this double-blind, randomized, placebo-controlled pharmaco-fMRI study, we examined the distinct neurobiological effects of an ayahuasca-inspired formulation containing DMT and harmine administered in conjunction with mindfulness-based practice in experienced meditation practitioners. Our goal was to explore both the individual and synergistic impacts of combining a 3-day meditation retreat with a psychedelic intervention versus placebo on brain function. We used several large-scale connectivity metrics, including within- and between-network connectivity, network and global connectivity, and cortical gradient analyses to assess these subacute effects following the retreat. Between-group comparisons revealed that participants in the DMT-harmine group showed increased FC between visual (VIS) and regions in salience (SAL) networks. Within-group comparisons further highlighted that the DMT-harmine group exhibited increased FC between DAN and regions in VIS, while the placebo group showed decreased FC between several RSNs. Although the within- and between-network results did not survive FDR correction, the observed trends were supported by the network connectivity analysis, which provided a more fine-grained, spatially sensitive perspective. These findings underscore the distinct neurobiological pathways engaged by meditation practice on placebo versus psychedelic-augmented meditation, contributing to a growing understanding of their potentially complementary and synergistic effects on brain function.

### 4.1. Meditation practice mainly decouples RSNs post-retreat

Our findings indicate that meditation alone induces subacute changes in several connectivity metrics, consistent with prior research. However, it is important to note that meditation studies usually do not include placebo administration, which may itself influence the subjective meditation experience and neural outcomes. Specifically, FC between multiple networks in the placebo group decreased, ranging from lower-order sensory regions (e.g., VIS) to higher-order attention and transmodal networks (e.g., SAL, DAN, FPN, and DMN). These effects remained consistent after applying GSR.

Our results are in line with previous findings (Ganesan et al., 2023; Hasenkamp & Barsalou, 2012a; Kilpatrick et al., 2011; Sezer et al., 2022). For instance, focused attention meditation has been associated with reduced connectivity across multiple brain regions (Ganesan et al., 2023), mirroring the connectivity reductions we observed in the placebo group post-retreat. Decreased VIS-SAL connectivity may reflect enhanced self-awareness (Sezer et al., 2022) or increased attentional control (Hasenkamp & Barsalou, 2012b; Kilpatrick et al., 2011), while increased FPN-DMN connectivity has also been linked to improved attentional regulation during meditation (Sezer et al., 2022).

However, some discrepancies with our findings remain; for example, an 8-day meditation retreat reported increased connectivity between SAL-DMN in trained meditators (Vishnubhotla et al., 2021) or other studies reported increased FPN-DMN FC after meditation training (Brewer et al., 2011; Kral et al., 2019). Differences in methodology, participant experience, meditation techniques, the use of a placebo, or retreat duration may explain such variations.

### 4.2. Effects of psychedelic-augmented meditation

The DMT-harmine administration led to trend-level increases in network integrity in the VIS, and increased between VIS–DAN connectivity. These effects contrasted with those observed in the placebo group, where connectivity generally decreased. A prior fMRI study investigating the combined effects of psilocybin during a 5-day meditation retreat found both increased and decreased FC in the DMN during open-awareness meditation in the psilocybin group (Smigielski et al., 2019). Despite the comparable design, these results were not replicated herein, perhaps due to several differences such as the psychedelic compound used or the analytical focus (i.e., DMN only vs. all Yeo networks).

Previous studies on the subacute effects of ayahuasca reported increased FC within SAL, both decreases (Pasquini et al., 2020) and increases in within-DMN FC with positive correlations to self-compassion measures (Sampedro et al., 2017), and increased FC between SAL-DMN (Pasquini et al., 2020). Decreased between-network FC between VIS and ACC has also been reported (Sampedro et al., 2017). While these studies also involved DMT and harmine administration, direct comparisons with our findings are complicated by differences in dosing and formulation. In our study, DMT and harmine were administered at a fixed 1:1 ratio (120 mg each over 90 minutes), whereas ayahuasca preparations in (Pasquini et al., 2020) and (Sampedro et al., 2017) involved a lower DMT-to-harmine ratio, with harmine doses approximately comparable to ours. From a pharmacological standpoint, this suggests that our formulation may have elicited stronger psychoactive effects (Egger et al., 2025). However, this was not clearly reflected in the observed connectivity patterns, which may indicate the important modulatory role of non-pharmacological factors such as the setting (meditation group retreat vs single-person study settings). For a more detailed discussion of pharmacological differences and similarities between ayahuasca and our DMT-harmine formulation, see Egger et al. (2024). Notably, previous studies highlight even longer-term effects of psilocybin. For example, Siegel and colleagues (2024) reported persistent reductions in functional connectivity between the anterior hippocampus and DMN in healthy volunteers, effects that endured for several weeks post-administration. Further structural evidence comes from a recent preprint, which investigated psilocybin’s long-term impact on measures like diffusion tensor imaging (DTI); it indicated decreased axial diffusivity in prefrontal-subcortical tracts, interpreted by the authors as potential pruning within a neuroplastic context (Lyons et al., 2024). These findings, underscoring prolonged neuroplasticity, are also critically important clinically. Indeed, psilocybin therapy has been shown to alter dynamic connectivity between the anterior and posterior cingulate cortices in patients with major depression, with these changes correlating with alterations in specific metabolites measured by spectroscopy. Moreover, evidence points to reduced between-network connectivity (i.e., modularity) persisting for weeks after psilocybin treatment in patients with treatment-resistant depression (TRD), a change that correlated with clinical improvement (Daws et al., 2022).

Beyond subacute effects, increased FC in the primary visual cortex (V1) during acute ayahuasca intake has been shown previously during a mental imagery task (De Araujo et al., 2012). Together, these findings suggest that ayahuasca alters the activity and connectivity of visual, salience, and DMN areas in healthy individuals. These previous reports are partially supported by our findings for the DMT-harmine formulation combined with mindfulness practice, which appeared to specifically alter VIS connectivity, both within-network and between visual areas and attentional networks (i.e., DAN).

Conversely, the placebo group showed reduced SAL-VIS FC, consistent with prior findings linking this to increased attentional focus and diminished irrelevant visual input during rest in experienced meditators (Hasenkamp & Barsalou, 2012b; Kilpatrick et al., 2011). Comparable patterns have also been observed following mindfulness-based cognitive therapy, where decreased SAL-VIS FC during rumination correlated with increased interoceptive awareness (van der Velden et al., 2023).

Contrary to our findings, acute intravenous DMT administration has been shown to reduce network integrity within the VIS and decrease segregation between VIS-FPN (Timmermann et al., 2023). However, our observed increase in voxel-wise FC between VIS and attentional networks may still be functionally related, as these networks, along with FPN, collectively facilitate attentional resource allocation (Menon, 2011; Vossel et al., 2014). As the previous study focused on acute drug effects versus placebo, direct comparisons are challenging.

### 4.3. Synergistic effects of meditation + DMT-harmine

Our findings revealed consistent group differences in FC, notably increased VIS-SAL connectivity in the DMT-harmine group. This result was robust across analyses comparing post-retreat scans and pre-to-post changes, with similar patterns emerging. Including GSR did not alter this primary finding, though it revealed additional group differences.

The increased VIS-SAL connectivity may reflect stronger engagement of visual and attentional processes during the psychedelic experience in participants receiving DMT-harmine, with residual effects detectable two days post-intervention. Another plausible mechanism involves heightened mental imagery induced by DMT-harmine during the retreat, potentially manifesting post-retreat at rest as visual recollections or “visions in reverse”, where information flows top-down rather than bottom-up, as in typical perception. In support, neutral or pleasant visual phenomena have also been reported within a week of LSD or psilocybin administration in ∼10% of participants (Müller et al., 2022). The SAL’s role in detecting and integrating sensory, emotional, and memory-related input (Schimmelpfennig et al., 2023) supports the idea that DMT-harmine facilitates the integration of visual and emotional information — an effect that may extend into the subacute phase. Relatedly, acute DMT has also been shown to increase connectivity between regions involved in visual, emotional, and associative processing, including the amygdala, supramarginal gyrus, and orbitofrontal cortex (Soares et al., 2024). Future studies could further investigate the relationship between the VIS and SAL by quantifying the effective connectivity to assess the directionality between these brain regions.

Additionally, this VIS-SAL cluster showed a significant interaction effect with mystical experience intensity (MEQ) and mindfulness (TMS) scores assessed on the day of drug administration. Interestingly, post-hoc analyses revealed a positive correlation between MEQ scores and SAL-VIS FC in the placebo group, but no significant correlations were found in the DMT-harmine group, despite a significant increase in MEQ scores in the latter (Meling et al., 2024). This suggests that the relationship between SAL-VIS FC and strength of the mystical experience and mindfulness might follow an inverse U-shaped curve, where neither too much nor too little connectivity is optimal.

Interestingly, this increased SAL-VIS FC differs from subacute changes observed after ayahuasca without meditation in a previous study, reporting increased SAL integrity and decreased SAL-DMN segregation (Pasquini et al., 2020). Sampedro et al. also reported both increased and decreased FC between DMN, VIS, and SAL regions (Sampedro et al., 2017), underscoring the importance of these networks in post-ayahuasca or DMT-harmine-induced changes.

In sum, our findings showed increased VIS integrity and enhanced VIS-DAN and VIS-SAL connectivity, diverging from our original hypotheses of increased within-SAL FC or stronger VIS-DMN and SAL-DMN coupling. These hypotheses were based on studies with different drug formulations, sample characteristics, and setting variables. For instance, prior meditation retreat studies have reported SAL-DMN connectivity increases (Vishnubhotla et al., 2021). Differences in retreat duration or other time-dependent variables may explain such discrepant findings.

From a broader perspective, the current results highlight neurobiological distinctions between the effects observed in meditation versus psychedelic-augmented meditation. While the meditation retreat with placebo mainly led to decreased FC between several RSNs that more closely reflect established neuroimaging findings of meditation alone (Hasenkamp & Barsalou, 2012a; Kilpatrick et al., 2011; Moujaes et al., 2024; Sezer et al., 2022; Vishnubhotla et al., 2021), psychedelic-augmented meditation mainly resulted in increased FC within the visual network and between visual and attention-related networks. Supporting this distinction, a recent study found that FC patterns were similar between acute psilocybin and LSD administration but differed when compared to meditation practiced in the scanner (Moujaes et al., 2024). While this distinction may not be entirely unexpected, it underscores an important point: Combining psychedelics with mindfulness-based interventions may offer complementary benefits in therapeutic settings, potentially enhancing both visual processing and attentional control.

The persistence of VIS-SAL connectivity changes two days post-administration indicates a potential window of opportunity for therapeutic interventions such as visual imagery exercises or to explore emotionally salient aspects of the experience in a mindfulness-oriented way.

Both psychedelics and mindfulness-based practices have independently demonstrated efficacy in alleviating symptoms of various affective disorders and warrant further exploration in clinical trials (Andersen et al., 2021; Milliere et al., 2018; Sezer et al., 2022). A potential mechanism of synergy lies in the increased awareness and mindfulness cultivated through meditation paired with the capacity of psychedelics to induce profound alterations in consciousness. This combination may be particularly beneficial during psychedelic therapy sessions, where the depth of the psychedelic experience has been correlated with therapeutic outcomes [79].

### 4.4. Disruption of cortical hierarchy: an acute psychedelic phenomenon?

A novel aspect of our study is the examination of cortical gradients and gradient dispersion in the subacute phase following psychedelic administration. The human principal cortical gradient, extending from the lower-order sensorimotor regions to the higher-order transmodal association cortex, serves as a fundamental organizational axis of the brain [69], whereas the second and third cortical gradients typically represent axes from the visual to insular cortex and from visual to areas covered by the FPN (Bethlehem et al., 2020; Girn et al., 2022), which is also evident in our data.

Recent neuroimaging studies suggest that altered states of consciousness, such as those induced by psychedelics like psilocybin, LSD, and DMT, acutely disrupt cortical gradient hierarchy (Girn et al., 2022; Timmermann et al., 2023). Psychedelics appear to reduce the differentiation of the evolutionarily younger, transmodal association cortex from lower-level sensory regions, an effect captured by a contraction along the principal cortical gradient (gradient 1). Beyond this, psychedelics also diminish differentiation along gradient 2, which separates visual regions from the insula and somatomotor cortices, and gradient 3, which distinguishes visual areas from FPN regions—suggesting a broad disruption of cortical functional organization (Girn et al., 2022; Timmermann et al., 2023).

This “collapse” of gradient axes aligns with the immersive, boundary-dissolving experiences commonly reported during acute psychedelic states and likely reflects a transient breakdown of established hierarchical organization in the brain. However, in our study, we found no evidence of such a collapse in cortical gradient architecture two days after the DMT-harmine or placebo retreat experience. This absence of residual flattening in the cortical hierarchy suggests that these alterations are acute and reversible, normalizing once the drug effects subside. It is also worth noting that our preprocessing pipeline employed a relatively stringent denoising strategy, which may have reduced sensitivity to subtle lasting changes in cortical gradient architecture. To further assess post-acute reorganization, we examined multi-dimensional gradient dispersion—a measure of how functionally distributed or spread cortical regions are within the gradient space (Bethlehem et al., 2020; Pasquini et al., 2023). While gradient dispersion has been shown to increase with aging, reflecting a dedifferentiation of functional brain organization, and to decrease in major depressive disorder (Pasquini et al., 2020), we observed no significant differences between groups or timepoints in our dataset. This further supports the notion that large-scale cortical functional architecture returns to baseline following the acute psychedelic state.

### 4.5. Limitations

Our study has several limitations that should be considered when interpreting the findings. A main issue in determining the synergistic effects of meditation and psychedelic intervention is the absence of both an intervention-free control group and a meditation-free psychedelic group. To fully understand the unique synergy that emerges when combining meditation and psychedelics, it would also be necessary to include a group receiving only the pharmacological intervention, without any meditation practice. Second, our study design included only subacute scans. While this adds to the currently limited literature of subacute effects of ayahuasca, or ayahuasca-inspired formulations (Pasquini et al., 2020; Sampedro et al., 2017), future studies incorporating both acute and subacute timepoints would provide a more complete understanding of these effects. Another limitation is the blinding efficacy, as most participants (>80%) recognized whether they were in the active or placebo group (for further details see (Meling et al., 2024)). However, we note that blinding is a ubiquitous problem in psychedelic research. The FDA has recently made some suggestions to overcome this issue, but a final solution has not yet been reached (Aday et al., 2025; *Psychedelic Drugs: Considerations for Clinical Investigations Guidance for Industry*, 2023). We prioritized a pharmacologically inert placebo to optimize neuroimaging contrasts, despite reduced blinding efficacy. Active or low-dose controls may better preserve blinding but risk confounding drug-specific neural effects. The specific study sample, primarily white and experienced meditators, limits the generalizability of our findings to broader populations. Notably, experienced meditators actively train to enter altered states of consciousness and may be more accustomed to such experiences than the general population. This familiarity might reduce their susceptibility to the perturbing effects of psychedelics, potentially contributing to the absence of strong effects observed two days after the meditation retreat in both treatment groups. Additionally, the group setting during the meditation retreat may have influenced participants’ experiences. Emphasis was placed on facilitating an introspective and meditative setting, but some minor degree of indirect interactions/influences between participants cannot be ruled out. The DMT-harmine and placebo groups were placed together, which could have led to an unintentional blending of experiences across conditions, potentially impacting connectivity outcomes and subjective experiences. Also, the study was conducted in two separate retreats, which had the same structure, but the two sessions could have influenced the outcomes of individual participants. Finally, the smoothing estimates for the network connectivity analysis were on the lower side with around 4–4.5 mm in each direction.

### 4.6. Conclusion

In conclusion, our findings reveal that the combined effects of DMT-harmine and meditation differ from meditation alone, with each intervention producing distinct functional connectivity changes that reflect their unique neurobiological mechanisms. Enhanced functional connectivity in visual and attention-related areas following a psychedelic-enhanced mindfulness retreat complements previous studies on psilocybin and ayahuasca in the context of mindfulness practices. The absence of cortical gradient disruption two days post-retreat highlights that the reorganization observed during acute psychedelic states is transient, emphasizing the brain’s ability to return to its hierarchical organization after the psychedelic experience subsides. These results suggest that psychedelic-augmented meditation may promote a more integrated brain state than meditation alone, potentially enhancing the depth and meaningfulness of subjective experiences.

## Data Availability

Imaging data related to this project will be made available in an online repository. Additional information and code is available upon reasonable request.

## Acknowledgements

We thank Jovin Müller, Javier Jareño Redondo for their medical support and screening of participants, Helena D Aicher, Joëlle Dornbierer, Elijah Temperli, Emilia A Vasella, Luzia Caflisch, David J Pfeiffer, Jonas TT Schlomberg, John W Smallridge, Alexandra Hempe and Sara Romer for their support in various aspects related to conducting this study, e.g. screening of participants, data collection and preparation. We additionally thank Matthias Hirsch-Hoffmann for his technical expertise and IT support in setting up the computational infrastructure necessary for our neuroimaging analyses, Peter Gasser and Karim Tissira for their medical support during the study interventions, Vanja Palmers and Miguel Guldimann for guiding the meditation study retreats, the Felsentor Foundation for hosting the meditation study retreats, and Reconnect Labs for providing the study drugs. Lastly, we thank the participants of this study.

This work was supported by the BIAL Foundation (No. 333/20), the Reconnect Foundation, and a European Varela Award from Mind and Life Europe (#2021EVA-Scheidegger, Milan). Any views, findings, conclusions, or recommendations expressed in this article do not necessarily reflect those of the funders.

## Author Contributions

**Klemens Egger:** Conceptualization, Data curation, Formal analysis, Investigation, Project administration, Visualization, Writing – Original draft, Writing – Review and editing. **Daniel Meling:** Conceptualization, Investigation, Project Administration, Writing – Review and editing. **Firuze Polat:** Investigation. **Erich Seifritz:** Resources. **Mihai Avram:** Methodology, Supervision, Writing – Review and editing. **Milan Scheidegger:** Funding Acquisition, Conceptualization, Methodology, Supervision, Writing – Review and editing.

## References

Aday, J. S., Simonsson, O., Schindler, E. A. D., & D’Souza, D. C. (2025). Addressing blinding in classic psychedelic studies with innovative active placebos. International Journal of Neuropsychopharmacology, 28(4). 10.1093/ijnp/pyaf023

Andersen, K. A. A., Carhart-Harris, R., Nutt, D. J., & Erritzoe, D. (2021). Therapeutic effects of classic serotonergic psychedelics: A systematic review of modern-era clinical studies. Acta Psychiatr Scand, 143(2), 101–118. 10.1111/acps.13249

Avram, M., Fortea, L., Wollner, L., Coenen, R., Korda, A., Rogg, H., Holze, F., Vizeli, P., Ley, L., Radua, J., Müller, F., Liechti, M. E., & Borgwardt, S. (2024). Large-scale brain connectivity changes following the administration of lysergic acid diethylamide, d-amphetamine, and 3,4-methylenedioxyamphetamine. Molecular Psychiatry. 10.1038/s41380-024-02734-y

Barrett, F. S., Johnson, M. W., & Griffiths, R. R. (2015). Validation of the revised Mystical Experience Questionnaire in experimental sessions with psilocybin. Journal of Psychopharmacology, 29(11), 1182–1190. 10.1177/0269881115609019

Behzadi, Y., Restom, K., Liau, J., & Liu, T. T. (2007). A component based noise correction method (CompCor) for BOLD and perfusion based fMRI. NeuroImage, 37(1), 90–101. 10.1016/J.NEUROIMAGE.2007.04.042

Bethlehem, R. A. I., Paquola, C., Seidlitz, J., Ronan, L., Bernhardt, B., Consortium, C.-C., & Tsvetanov, K. A. (2020). Dispersion of functional gradients across the adult lifespan. NeuroImage, 222, 117299. 10.1016/j.neuroimage.2020.117299

Brewer, J. A., Worhunsky, P. D., Gray, J. R., Tang, Y.-Y., Weber, J., & Kober, H. (2011). Meditation experience is associated with differences in default mode network activity and connectivity. Proceedings of the National Academy of Sciences, 108(50), 20254–20259. 10.1073/pnas.1112029108

Carhart-Harris, R. L., Leech, R., Hellyer, P. J., Shanahan, M., Feilding, A., Tagliazucchi, E., Chialvo, D. R., & Nutt, D. (2014). The entropic brain: a theory of conscious states informed by neuroimaging research with psychedelic drugs. Front Hum Neurosci, 8, 20. 10.3389/fnhum.2014.00020

Chumbley, J., Worsley, K., Flandin, G., & Friston, K. (2010). Topological FDR for neuroimaging. NeuroImage, 49(4), 3057–3064. 10.1016/j.neuroimage.2009.10.090

Coifman, R. R., Lafon, S., Lee, A. B., Maggioni, M., Nadler, B., Warner, F., & Zucker, S. W. (2005). Geometric diffusions as a tool for harmonic analysis and structure definition of data: Diffusion maps. Proceedings of the National Academy of Sciences, 102(21), 7426–7431. 10.1073/pnas.0500334102

Dahl, C. J., Wilson-Mendenhall, C. D., & Davidson, R. J. (2020). The plasticity of well-being: A training-based framework for the cultivation of human flourishing. Proceedings of the National Academy of Sciences, 117(51), 32197–32206. 10.1073/pnas.2014859117

Davidson, R. J. (2021). Mindfulness and More: Toward a Science of Human Flourishing. Psychosomatic Medicine, 83(6), 665–668. 10.1097/PSY.0000000000000960

Daws, R. E., Timmermann, C., Giribaldi, B., Sexton, J. D., Wall, M. B., Erritzoe, D., Roseman, L., Nutt, D., & Carhart-Harris, R. (2022). Increased global integration in the brain after psilocybin therapy for depression. Nature Medicine, 28(4), 844–851. 10.1038/s41591-022-01744-z

De Araujo, D. B., Ribeiro, S., Cecchi, G. A., Carvalho, F. M., Sanchez, T. A., Pinto, J. P., de Martinis, B. S., Crippa, J. A., Hallak, J. E. C., & Santos, A. C. (2012). Seeing with the eyes shut: neural basis of enhanced imagery following Ayahuasca ingestion. Human Brain Mapping, 33(11), 2550–2560. 10.1002/HBM.21381

Dominguez-Clave, E., Soler, J., Elices, M., Pascual, J. C., Alvarez, E., de la Fuente Revenga, M., Friedlander, P., Feilding, A., & Riba, J. (2016). Ayahuasca: Pharmacology, neuroscience and therapeutic potential. Brain Res Bull, 126(Pt 1), 89–101. 10.1016/j.brainresbull.2016.03.002

Dornbierer, D. A., Marten, L., Mueller, J., Aicher, H. D., Mueller, M. J., Boxler, M., Kometer, M., Kosanic, D., von Rotz, R., Puchkov, M., Kraemer, T., Landolt, H.-P., Seifritz, E., & Scheidegger, M. (2023). Overcoming the clinical challenges of traditional ayahuasca: a first-in-human trial exploring novel routes of administration of N,N-Dimethyltryptamine and harmine. Frontiers in Pharmacology, 14. 10.3389/fphar.2023.1246892

Egger, K., Aicher, H. D., Cumming, P., & Scheidegger, M. (2024). Neurobiological research on N,N-dimethyltryptamine (DMT) and its potentiation by monoamine oxidase (MAO) inhibition: from ayahuasca to synthetic combinations of DMT and MAO inhibitors. Cellular and Molecular Life Sciences, 81(1), 1–26. 10.1007/S00018-024-05353-6

Egger, K., Jareño Redondo, J., Müller, J., Dornbierer, J., Smallridge, J., Aicher, H. D., Meling, D., Müller, P., Kost, J., Puchkov, M., Äbelö, A., Seifritz, E., Quednow, B. B., von Rotz, R., Scheidegger, M., & Dornbierer, D. A. (2025). Examining the pharmacokinetic and pharmacodynamic interaction of N,N-dimethyltryptamine and harmine in healthy volunteers: ? factorial dose-escalation study. Biomedicine & Pharmacotherapy, 184, 117908. 10.1016/J.BIOPHA.2025.117908

Esteban, O., Markiewicz, C. J., Blair, R. W., Moodie, C. A., Isik, A. I., Erramuzpe, A., Kent, J. D., Goncalves, M., DuPre, E., Snyder, M., Oya, H., Ghosh, S. S., Wright, J., Durnez, J., Poldrack, R. A., & Gorgolewski, K. J. (2019). fMRIPrep: a robust preprocessing pipeline for functional MRI. Nature Methods, 16(1), 111–116. 10.1038/s41592-018-0235-4

Esteban, O., Markiewicz, C. J., Goncalves, M., Provins, C., Kent, J. D., DuPre, E., Salo, T., Ciric, R., Pinsard, B., Blair, R. W., Poldrack, R. A., & Gorgolewski, K. J. (2023). fMRIPrep 23.0.2. Zenodo. 10.5281/zenodo.7863421

Falchi-Carvalho, M., Barros, H., Bolcont, R., Laborde, S., Wießner, I., Ruschi B. Silva, S., Montanini, D., Barbosa, D. C., Teixeira, E., Florence-Vilela, R., Almeida, R., de Macedo, R. K. A., Arichelle, F., Pantrigo, É.J., Costa-Macedo, J. V., Arcoverde, E., Galvão-Coelho, N., Araujo, D. B., & Palhano-Fontes, F. (2025). The Antidepressant Effects of Vaporized N, N -Dimethyltryptamine: An Open-Label Pilot Trial in Treatment-Resistant Depression. Psychedelic Medicine, 3(1), 48–52. 10.1089/psymed.2024.0002

Friston, K. J., Williams, S., Howard, R., Frackowiak, R. S. J., & Turner, R. (1996). Movement-Related effects in fMRI time-series. Magnetic Resonance in Medicine, 35(3), 346–355. 10.1002/mrm.1910350312

Ganesan, S. A., Moffat, B., Van Dam, N. T., Lorenzetti, V., & Zalesky, A. (2023). Meditation attenuates default-mode activity: A pilot study using ultra-high field 7 Tesla MRI. Brain Research Bulletin, 203, 110766. 10.1016/j.brainresbull.2023.110766

Girn, M., Roseman, L., Bernhardt, B., Smallwood, J., Carhart-Harris, R., & Nathan Spreng, R. (2022). Serotonergic psychedelic drugs LSD and psilocybin reduce the hierarchical differentiation of unimodal and transmodal cortex. NeuroImage, 256, 119220. 10.1016/j.neuroimage.2022.119220

Goldberg, S. B., Tucker, R. P., Greene, P. A., Davidson, R. J., Wampold, B. E., Kearney, D. J., & Simpson, T. L. (2018). Mindfulness-based interventions for psychiatric disorders: A systematic review and meta-analysis. Clinical Psychology Review, 59, 52–60. 10.1016/j.cpr.2017.10.011

Gordon, E. M., Laumann, T. O., Adeyemo, B., Huckins, J. F., Kelley, W. M., & Petersen, S. E. (2016). Generation and Evaluation of a Cortical Area Parcellation from Resting-State Correlations. Cerebral Cortex, 26(1), 288–303. 10.1093/cercor/bhu239

Griffiths, R. R., Johnson, M. W., Richards, W. A., Richards, B. D., Jesse, R., MacLean, K. A., Barrett, F. S., Cosimano, M. P., & Klinedinst, M. A. (2018). Psilocybin-occasioned mystical-type experience in combination with meditation and other spiritual practices produces enduring positive changes in psychological functioning and in trait measures of prosocial attitudes and behaviors. Journal of Psychopharmacology (Oxford, England), 32(1), 49–69. 10.1177/0269881117731279

Gu, J., Baer, R., Cavanagh, K., Kuyken, W., & Strauss, C. (2020). Development and Psychometric Properties of the Sussex-Oxford Compassion Scales (SOCS). Assessment, 27(1), 3–20. 10.1177/1073191119860911

Hallquist, M. N., Hwang, K., & Luna, B. (2013). The nuisance of nuisance regression: Spectral misspecification in a common approach to resting-state fMRI preprocessing reintroduces noise and obscures functional connectivity. NeuroImage, 82, 208–225. 10.1016/j.neuroimage.2013.05.116

Hasenkamp, W., & Barsalou, L. W. (2012a). Effects of meditation experience on functional connectivity of distributed brain networks. Front Hum Neurosci, 6, 38. 10.3389/fnhum.2012.00038

Hasenkamp, W., & Barsalou, L. W. (2012b). Effects of Meditation Experience on Functional Connectivity of Distributed Brain Networks. Frontiers in Human Neuroscience, 6. 10.3389/fnhum.2012.00038

Kilpatrick, L. A., Suyenobu, B. Y., Smith, S. R., Bueller, J. A., Goodman, T., Creswell, J. D., Tillisch, K., Mayer, E. A., & Naliboff, B. D. (2011). Impact of mindfulness-based stress reduction training on intrinsic brain connectivity. NeuroImage, 56(1), 290– 298. 10.1016/j.neuroimage.2011.02.034

Ko, K., Knight, G., Rucker, J. J., & Cleare, A. J. (2022). Psychedelics, Mystical Experience, and Therapeutic Efficacy: A Systematic Review. Frontiers in Psychiatry, v13. 10.3389/fpsyt.2022.917199

Kost, J., Huwyler, J., & Puchkov, M. (2023). Calcium Phosphate Microcapsules as Multifunctional Drug Delivery Devices. Advanced Functional Materials, 33(38). 10.1002/adfm.202303333

Kral, T. R. A., Imhoff-Smith, T., Dean, D. C., Grupe, D., Adluru, N., Patsenko, E., Mumford, J. A., Goldman, R., Rosenkranz, M. A., & Davidson, R. J. (2019). Mindfulness-Based Stress Reduction-related changes in posterior cingulate resting brain connectivity. Social Cognitive and Affective Neuroscience, 14(7), 777– 787. 10.1093/scan/nsz050

Larivière, S., Bayrak, Ş., Vos de Wael, R., Benkarim, O., Herholz, P., Rodriguez-Cruces, R., Paquola, C., Hong, S.-J., Misic, B., Evans, A. C., Valk, S. L., & Bernhardt, B. C. (2023). BrainStat: A toolbox for brain-wide statistics and multimodal feature associations. NeuroImage, 266, 119807. 10.1016/j.neuroimage.2022.119807

Lau, M. A., Bishop, S. R., Segal, Z. V., Buis, T., Anderson, N. D., Carlson, L., Shapiro, S., Carmody, J., Abbey, S., & Devins, G. (2006). The toronto mindfulness scale: Development and validation. Journal of Clinical Psychology, 62(12), 1445–1467. 10.1002/jclp.20326

Lebedev, A. V, Kaelen, M., Lövdén, M., Nilsson, J., Feilding, A., Nutt, D. J., & Carhart-Harris, R. L. (2016). LSD-induced entropic brain activity predicts subsequent personality change. Hum Brain Mapp, 37(9), 3203–3213. 10.1002/hbm.23234

Lyons, T., Spriggs, M., Kerkelä, L., Rosas, F., Roseman, L., Mediano, P., Timmermann, C., Oestreich, L., Pagni, B., Zeifman, R., Hampshire, A., Trender, W., Douglass, H., Girn, M., Godfrey, K., Kettner, H., Sharif, F., Espasiano, L., Gazzaley, A., … Carhart-Harris, R. (2024). Human brain changes after first psilocybin use. BioRxiv. 10.1101/2024.10.11.617955

Madsen, M. K., Stenbæk, D. S., Arvidsson, A., Armand, S., Marstrand-Joergensen, M. R., Johansen, S. S., Linnet, K., Ozenne, B., Knudsen, G. M., & Fisher, P. M. (2021). Psilocybin-induced changes in brain network integrity and segregation correlate with plasma psilocin level and psychedelic experience. European Neuropsychopharmacology, 50, 121–132. 10.1016/j.euroneuro.2021.06.001

Maia, L. O., Daldegan-Bueno, D., Wießner, I., Araujo, D. B., & Tófoli, L. F. (2023). Ayahuasca’s therapeutic potential: What we know – and what not. European Neuropsychopharmacology, 66, 45–61. 10.1016/j.euroneuro.2022.10.008

Mallaroni, P., Mason, N. L., Kloft, L., Reckweg, J. T., van Oorsouw, K., Toennes, S. W., Tolle, H. M., Amico, E., & Ramaekers, J. G. (2024). Shared functional connectome fingerprints following ritualistic ayahuasca intake. NeuroImage, 285, 120480. 10.1016/j.neuroimage.2023.120480

Margulies, D. S., Ghosh, S. S., Goulas, A., Falkiewicz, M., Huntenburg, J. M., Langs, G., Bezgin, G., Eickhoff, S. B., Castellanos, F. X., Petrides, M., Jefferies, E., & Smallwood, J. (2016). Situating the default-mode network along a principal gradient of macroscale cortical organization. Proceedings of the National Academy of Sciences, 113(44), 12574–12579. 10.1073/pnas.1608282113

McCulloch, D. E.-W., Knudsen, G. M., Barrett, F. S., Doss, M. K., Carhart-Harris, R. L., Rosas, F. E., Deco, G., Kringelbach, M. L., Preller, K. H., Ramaekers, J. G., Mason, N. L., Müller, F., & Fisher, P. M. (2022). Psychedelic resting-state neuroimaging: A review and perspective on balancing replication and novel analyses. Neuroscience & Biobehavioral Reviews, 138, 104689. 10.1016/j.neubiorev.2022.104689

Meling, D., Egger, K., Aicher, H. D., Jareño Redondo, J., Mueller, J., Dornbierer, J., Temperli, E., Vasella, E. A., Caflisch, L., Pfeiffer, D. J., Schlomberg, J. T., Smallridge, J. W., Dornbierer, D. A., & Scheidegger, M. (2024). Meditating on psychedelics. A randomized placebo-controlled study of DMT and harmine in a mindfulness retreat. Journal of Psychopharmacology. 10.1177/02698811241282637

Menon, V. (2011). Large-scale brain networks and psychopathology: a unifying triple network model. Trends in Cognitive Sciences, 15(10), 483–506. 10.1016/j.tics.2011.08.003

Milliere, R., Carhart-Harris, R. L., Roseman, L., Trautwein, F. M., & Berkovich-Ohana, A. (2018). Psychedelics, Meditation, and Self-Consciousness. Front Psychol, 9, 1475. 10.3389/fpsyg.2018.01475

Moujaes, F., Rieser, N. M., Phillips, C., de Matos, N. M. P., Brügger, M., Dürler, P., Smigielski, L., Stämpfli, P., Seifritz, E., Vollenweider, F. X., Anticevic, A., & Preller, K. H. (2024). Comparing Neural Correlates of Consciousness: From Psychedelics to Hypnosis and Meditation. Biological Psychiatry: Cognitive Neuroscience and Neuroimaging, 9(5), 533–543. 10.1016/j.bpsc.2023.07.003

Müller, F., Kraus, E., Holze, F., Becker, A., Ley, L., Schmid, Y., Vizeli, P., Liechti, M. E., & Borgwardt, S. (2022). Flashback phenomena after administration of LSD and psilocybin in controlled studies with healthy participants. Psychopharmacology, 239(6), 1933–1943. 10.1007/s00213-022-06066-z

Muttoni, S., Ardissino, M., & John, C. (2019). Classical psychedelics for the treatment of depression and anxiety: A systematic review. J Affect Disord, 258, 11–24. 10.1016/j.jad.2019.07.076

Nichols, D. E. (2016). Psychedelics. Pharmacol Rev, 68(2), 264–355. 10.1124/pr.115.011478

Nieto-Castanon, A. (2020). Handbook of functional connectivity Magnetic Resonance Imaging methods in CONN. Hilbert Press. 10.56441/hilbertpress.2207.6598

Nieto-Castanon, A., & Whitfield-Gabrieli, S. (2022). CONN functional connectivity toolbox: RRID SCR_009550, release 22. Hilbert Press. 10.56441/hilbertpress.2246.5840

Pasquini, L., Fryer, S. L., Eisendrath, S. J., Segal, Z. V., Lee, A. J., Brown, J. A., Saggar, M., & Mathalon, D. H. (2023). Dysfunctional Cortical Gradient Topography in Treatment-Resistant Major Depressive Disorder. Biological Psychiatry: Cognitive Neuroscience and Neuroimaging, 8(9), 928–939. 10.1016/j.bpsc.2022.10.009

Pasquini, L., Palhano-Fontes, F., & Araujo, D. B. (2020). Subacute effects of the psychedelic ayahuasca on the salience and default mode networks. J Psychopharmacol, 34(6), 623–635. 10.1177/0269881120909409

Peill, J. M., Trinci, K. E., Kettner, H., Mertens, L. J., Roseman, L., Timmermann, C., Rosas, F. E., Lyons, T., & Carhart-Harris, R. L. (2022). Validation of the Psychological Insight Scale: A new scale to assess psychological insight following a psychedelic experience. Journal of Psychopharmacology, 36(1), 31–45. 10.1177/02698811211066709

Power, J. D., Mitra, A., Laumann, T. O., Snyder, A. Z., Schlaggar, B. L., & Petersen, S. E. (2014). Methods to detect, characterize, and remove motion artifact in resting state fMRI. NeuroImage, 84, 320–341. 10.1016/J.NEUROIMAGE.2013.08.048

Preller, K. H., Burt, J. B., Ji, J. L., Schleifer, C. H., Adkinson, B. D., Stampfli, P., Seifritz, E., Repovs, G., Krystal, J. H., Murray, J. D., Vollenweider, F. X., & Anticevic, A. (2018). Changes in global and thalamic brain connectivity in LSD-induced altered states of consciousness are attributable to the 5-HT2A receptor. Elife, 7. 10.7554/eLife.35082

Preller, K. H., Duerler, P., Burt, J. B., Ji, J. L., Adkinson, B., Stämpfli, P., Seifritz, E., Repovš, G., Krystal, J. H., Murray, J. D., Anticevic, A., & Vollenweider, F. X. (2020). Psilocybin Induces Time-Dependent Changes in Global Functional Connectivity. Biological Psychiatry, 88(2), 197–207. 10.1016/j.biopsych.2019.12.027

Psychedelic Drugs: Considerations for Clinical Investigations Guidance for Industry. (2023, June). U.S. Department of Health and Human Services, Food and Drug Administration, Center for Drug Evaluation and Research (CDER). https://www.fda.gov/regulatory-information/search-fda-guidance-documents/psychedelic-drugs-considerations-clinical-investigations

Ramaekers, J. G., Mallaroni, P., Kloft, L., Reckweg, J. T., Toennes, S. W., van Oorsouw, K., & Mason, N. L. (2023). Altered State of Consciousness and Mental Imagery as a Function of N, N -dimethyltryptamine Concentration in Ritualistic Ayahuasca Users. Journal of Cognitive Neuroscience, 35(9), 1382–1393. 10.1162/jocn_a_02003

Ramaekers, J. G., Reckweg, J. T., & Mason, N. L. (2025). Benefits and Challenges of Ultra-Fast, Short-Acting Psychedelics in the Treatment of Depression. American Journal of Psychiatry, 182(1), 33–46. 10.1176/appi.ajp.20230890

Riba, J., McIlhenny, E. H., Valle, M., Bouso, J. C., & Barker, S. A. (2012). Metabolism and disposition of N,N-dimethyltryptamine and harmala alkaloids after oral administration of ayahuasca. Drug Test Anal, 4(7–8), 610–616. 10.1002/dta.1344

Romeo, B., Karila, L., Martelli, C., & Benyamina, A. (2020). Efficacy of psychedelic treatments on depressive symptoms: A meta-analysis. Journal of Psychopharmacology, 34(10), 1079–1085. 10.1177/0269881120919957

Rose, S., Zell, E., & Strickhouser, J. E. (2020). The Effect of Meditation on Health: a Metasynthesis of Randomized Controlled Trials. Mindfulness, 11(2), 507–516. 10.1007/s12671-019-01277-6

Roseman, L., Haijen, E., Idialu-Ikato, K., Kaelen, M., Watts, R., & Carhart-Harris, R. (2019). Emotional breakthrough and psychedelics: Validation of the Emotional Breakthrough Inventory. J Psychopharmacol, 33(9), 1076–1087. 10.1177/0269881119855974

Sampedro, F., de la Fuente Revenga, M., Valle, M., Roberto, N., Dominguez-Clave, E., Elices, M., Luna, L. E., Crippa, J. A. S., Hallak, J. E. C., de Araujo, D. B., Friedlander, P., Barker, S. A., Alvarez, E., Soler, J., Pascual, J. C., Feilding, A., & Riba, J. (2017). Assessing the Psychedelic ‘After-Glow’ in Ayahuasca Users: Post-Acute Neurometabolic and Functional Connectivity Changes Are Associated with Enhanced Mindfulness Capacities. Int J Neuropsychopharmacol, 20(9), 698–711. 10.1093/ijnp/pyx036

Schaefer, A., Kong, R., Gordon, E. M., Laumann, T. O., Zuo, X.-N., Holmes, A. J., Eickhoff, S. B., & Yeo, B. T. T. (2018). Local-Global Parcellation of the Human Cerebral Cortex from Intrinsic Functional Connectivity MRI. Cerebral Cortex, 28(9), 3095–3114. 10.1093/cercor/bhx179

Schimmelpfennig, J., Topczewski, J., Zajkowski, W., & Jankowiak-Siuda, K. (2023). The role of the salience network in cognitive and affective deficits. Frontiers in Human Neuroscience, 17, 1133367. 10.3389/fnhum.2023.1133367

Sezer, I., Pizzagalli, D. A., & Sacchet, M. D. (2022). Resting-state fMRI functional connectivity and mindfulness in clinical and non-clinical contexts: A review and synthesis. Neuroscience & Biobehavioral Reviews, 135, 104583. 10.1016/j.neubiorev.2022.104583

Siegel, J. S., Subramanian, S., Perry, D., Kay, B. P., Gordon, E. M., Laumann, T. O., Reneau, T. R., Metcalf, N. V., Chacko, R. V., Gratton, C., Horan, C., Krimmel, S. R., Shimony, J. S., Schweiger, J. A., Wong, D. F., Bender, D. A., Scheidter, K. M., Whiting, F. I., Padawer-Curry, J. A., … Dosenbach, N. U. F. (2024). Psilocybin desynchronizes the human brain. Nature, 632(8023), 131–138. 10.1038/s41586-024-07624-5

Singer, B., Meling, D., Hirsch-Hoffmann, M., Michels, L., Kometer, M., Smigielski, L., Dornbierer, D., Seifritz, E., Vollenweider, F. X., & Scheidegger, M. (2024). Psilocybin enhances insightfulness in meditation: a perspective on the global topology of brain imaging during meditation. Scientific Reports, 14(1), 7211. 10.1038/s41598-024-55726-x

Smigielski, L., Scheidegger, M., Kometer, M., & Vollenweider, F. X. (2019). Psilocybin-assisted mindfulness training modulates self-consciousness and brain default mode network connectivity with lasting effects. Neuroimage, 196, 207–215. 10.1016/j.neuroimage.2019.04.009

Soares, C., Lima, G., Pais, M. L., Teixeira, M., Cabral, C., & Castelo-Branco, M. (2024). Increased functional connectivity between brain regions involved in social cognition, emotion and affective-value in psychedelic states induced by N,N-Dimethyltryptamine (DMT). Frontiers in Pharmacology, 15. 10.3389/fphar.2024.1454628

Sydnor, V. J., Larsen, B., Bassett, D. S., Alexander-Bloch, A., Fair, D. A., Liston, C., Mackey, A. P., Milham, M. P., Pines, A., Roalf, D. R., Seidlitz, J., Xu, T., Raznahan, A., & Satterthwaite, T. D. (2021a). Neurodevelopment of the association cortices: Patterns, mechanisms, and implications for psychopathology. Neuron, 109(18), 2820–2846. 10.1016/j.neuron.2021.06.016

Sydnor, V. J., Larsen, B., Bassett, D. S., Alexander-Bloch, A., Fair, D. A., Liston, C., Mackey, A. P., Milham, M. P., Pines, A., Roalf, D. R., Seidlitz, J., Xu, T., Raznahan, A., & Satterthwaite, T. D. (2021b). Neurodevelopment of the association cortices: Patterns, mechanisms, and implications for psychopathology. Neuron, 109(18), 2820–2846. 10.1016/j.neuron.2021.06.016

Thomas, G., Lucas, P., Capler, N., Tupper, K., & Martin, G. (2013). Ayahuasca-Assisted Therapy for Addiction: Results from a Preliminary Observational Study in Canada. Current Drug Abuse Reviews, 6(1), 30–42. 10.2174/15733998113099990003

Timmermann, C., Roseman, L., Haridas, S., Rosas, F. E., Luan, L., Kettner, H., Martell, J., Erritzoe, D., Tagliazucchi, E., Pallavicini, C., Girn, M., Alamia, A., Leech, R., Nutt, D. J., & Carhart-Harris, R. L. (2023). Human brain effects of DMT assessed via EEG-fMRI. Proceedings of the National Academy of Sciences, 120(13). 10.1073/pnas.2218949120

Uthaug, M. V., Mason, N. L., Toennes, S. W., Reckweg, J. T., de Sousa Fernandes Perna, E.B., Kuypers, K. P. C., van Oorsouw, K., Riba, J., & Ramaekers, J. G. (2021). A placebo-controlled study of the effects of ayahuasca, set and setting on mental health of participants in ayahuasca group retreats. Psychopharmacology, 238(7), 1899–1910. 10.1007/S00213-021-05817-8

Van Dam, N. T., van Vugt, M. K., Vago, D. R., Schmalzl, L., Saron, C. D., Olendzki, A., Meissner, T., Lazar, S. W., Kerr, C. E., Gorchov, J., Fox, K. C. R., Field, B. A., Britton, W. B., Brefczynski-Lewis, J. A., & Meyer, D. E. (2018). Mind the Hype: A Critical Evaluation and Prescriptive Agenda for Research on Mindfulness and Meditation. Perspectives on Psychological Science, 13(1), 36–61. 10.1177/1745691617709589

van der Velden, A. M., Scholl, J., Elmholdt, E.-M., Fjorback, L. O., Harmer, C. J., Lazar, S. W., O’Toole, M. S., Smallwood, J., Roepstorff, A., & Kuyken, W. (2023). Mindfulness Training Changes Brain Dynamics During Depressive Rumination: A Randomized Controlled Trial. Biological Psychiatry, 93(3), 233–242. 10.1016/j.biopsych.2022.06.038

Vishnubhotla, R. V., Radhakrishnan, R., Kveraga, K., Deardorff, R., Ram, C., Pawale, D., Wu, Y.-C., Renschler, J., Subramaniam, B., & Sadhasivam, S. (2021). Advanced Meditation Alters Resting-State Brain Network Connectivity Correlating With Improved Mindfulness. Frontiers in Psychology, 12. 10.3389/fpsyg.2021.745344

Vollenweider, F. X., & Preller, K. H. (2020). Psychedelic drugs: neurobiology and potential for treatment of psychiatric disorders. Nat Rev Neurosci, 21(11), 611–624. 10.1038/s41583-020-0367-2

Vos de Wael, R., Benkarim, O., Paquola, C., Lariviere, S., Royer, J., Tavakol, S., Xu, T., Hong, S.-J., Langs, G., Valk, S., Misic, B., Milham, M., Margulies, D., Smallwood, J., & Bernhardt, B. C. (2020). BrainSpace: a toolbox for the analysis of macroscale gradients in neuroimaging and connectomics datasets. Communications Biology, 3(1), 103. 10.1038/s42003-020-0794-7

Vossel, S., Geng, J. J., & Fink, G. R. (2014). Dorsal and ventral attention systems: distinct neural circuits but collaborative roles. The Neuroscientist, 20(2), 150–159. 10.1177/1073858413494269

Whitfield-Gabrieli, S., & Nieto-Castanon, A. (2012). Conn: a functional connectivity toolbox for correlated and anticorrelated brain networks. Brain Connectivity, 2(3), 125–141. 10.1089/brain.2012.0073

Worsley, K. J., Marrett, S., Neelin, P., Vandal, A. C., Friston, K. J., & Evans, A. C. (1996). A unified statistical approach for determining significant signals in images of cerebral activation. Human Brain Mapping, 4(1), 58–73. 10.1002/(SICI)1097-0193(1996)4:1<58::AID-HBM4>3.0.CO;2-O

Yeo, B. T. T., Krienen, F. M., Chee, M. W. L., & Buckner, R. L. (2014). Estimates of segregation and overlap of functional connectivity networks in the human cerebral cortex. NeuroImage, 88, 212–227. 10.1016/j.neuroimage.2013.10.046

Yeo, B. T. T., Krienen, F. M., Sepulcre, J., Sabuncu, M. R., Lashkari, D., Hollinshead, M., Roffman, J. L., Smoller, J. W., Zöllei, L., Polimeni, J. R., Fischl, B., Liu, H., & Buckner, R. L. (2011). The organization of the human cerebral cortex estimated by intrinsic functional connectivity. Journal of Neurophysiology, 106(3), 1125–1165. 10.1152/jn.00338.2011

