## Supplementary Material for "Meditation, Psychedelics, and Brain Connectivity: A Randomised Controlled Resting-State fMRI Study of *N,N*-Dimethyltryptamine and Harmine in a Meditation Retreat"

### Suppl. Methods

#### Anatomical data preprocessing

Results included in this manuscript come from preprocessing performed using *fMRIPrep* 23.0.2 (Esteban et al., 2019, 2023), which is based on *Nipype* 1.8.6 (Esteban et al., 2022; Gorgolewski et al., 2011). The T1-weighted (T1w) image was corrected for intensity non-uniformity (INU) with *N4BiasFieldCorrection* (Tustison et al., 2010), distributed with ANTs 2.3.3 (Avants et al., 2008), and used as T1w-reference throughout the workflow. The T1w-reference was then skull-stripped with a *Nipype* implementation of the *antsBrainExtraction.sh* workflow (from ANTs), using OASIS30ANTs as target template. Brain tissue segmentation of cerebrospinal fluid (CSF), white-matter (WM) and gray-matter (GM) was performed on the brain-extracted T1w using *fast* (FSL 6.0.5.1:57b01774, (Zhang et al., 2001)). Brain surfaces were reconstructed using *recon-all* (FreeSurfer 7.3.2, (Dale et al., 1999)), and the brain mask estimated previously was refined with a custom variation of the method to reconcile ANTs-derived and FreeSurfer-derived segmentations of the cortical gray-matter of Mindboggle (Klein et al., 2017). Volume-based spatial normalization was performed through nonlinear registration with *antsRegistration* (ANTs 2.3.3), using brain-extracted versions of both T1w reference and the T1w template. The following template was selected for spatial normalization and accessed with *TemplateFlow* (23.0.0, (Ciric et al., 2022)): *ICBM 152 Nonlinear Asymmetrical template version 2009c* ((Fonov et al., 2009), TemplateFlow ID: MNI152NLin2009cAsym).

#### Functional data preprocessing

For each of the 2 BOLD runs per subject (pre- and post-intervention), the following preprocessing was performed. First, a reference volume and its skull-stripped version were generated using a custom methodology of *fMRIPrep*. Head-motion parameters with respect to the BOLD reference (transformation matrices, and six corresponding rotation and translation parameters) are estimated before any spatiotemporal filtering using *mcflirt* (FSL 6.0.5.1:57b01774, (Jenkinson et al., 2002)). BOLD runs were slice-time corrected to 0.884s (0.5 of slice acquisition range 0s-1.77s) using *3dTshift* from AFNI (Cox & Hyde, 1997). The BOLD time-series (including slice-timing correction) were resampled onto their original, native space by applying the transforms to correct for head-motion. These resampled BOLD time-series will be referred to as *preprocessed BOLD in original space*, or just *preprocessed BOLD*. The BOLD reference was then co-registered to the T1w reference using *bbregister* (FreeSurfer) which implements boundary-based registration (Greve & Fischl, 2009). Co-registration was configured with six degrees of freedom. Several confounding time-series were calculated based on the *preprocessed BOLD*: framewise displacement (FD), DVARS and three region-wise global signals. FD was computed using two formulations following Power (absolute sum of relative motions, (Power et al., 2014)) and Jenkinson (relative root mean square displacement between affines, (Jenkinson et al., 2002)). FD and DVARS are calculated for each functional run, both using their implementations in *Nipype* (following the definitions by Power et al. (Power et al., 2014)). The three global signals are extracted within the CSF, the WM, and the whole-brain masks. Additionally, a set of physiological regressors were extracted to allow for component-based noise correction (*CompCor*, (Behzadi et al., 2007)). Principal components are estimated after high-pass filtering the *preprocessed BOLD* time-series (using a discrete cosine filter with 128s cut-off) for anatomical *CompCor* (aCompCor). For aCompCor, three probabilistic masks (CSF, WM and combined CSF+WM) are generated in anatomical space. The implementation differs from that of Behzadi et al. (Behzadi et al., 2007) in that instead of eroding the masks by

2 pixels on BOLD space, a mask of pixels that likely contain a volume fraction of GM is subtracted from the aCompCor masks. This mask is obtained by dilating a GM mask extracted from the FreeSurfer's *aseg* segmentation, and it ensures components are not extracted from voxels containing a minimal fraction of GM. Finally, these masks are resampled into BOLD space and binarized by thresholding at 0.99 (as in the original implementation). Components are also calculated separately within the WM and CSF masks. For each aCompCor decomposition, the *k* components with the largest singular values are retained, such that the retained components' time series are sufficient to explain 50 percent of variance across the nuisance mask (CSF, WM, combined, or temporal). The remaining components are dropped from consideration. The head-motion estimates calculated in the correction step were also placed within the corresponding confounds file. The confound time series derived from head motion estimates and global signals were expanded with the inclusion of temporal derivatives and quadratic terms for each (Satterthwaite et al., 2013). Frames that exceeded a threshold of 0.5 mm FD or 1.5 standardized DVARS were annotated as motion outliers. The BOLD time-series were resampled into standard space, generating a *preprocessed BOLD run in MNI152NLin2009cAsym space*. First, a reference volume and its skull-stripped version were generated using a custom methodology of *fMRIPrep*. All resamplings can be performed with *a single interpolation step* by composing all the pertinent transformations (i.e., head-motion transformation matrices, susceptibility distortion correction when available, and co-registrations to anatomical and output spaces). Gridded (volumetric) resamplings were performed using *antsApplyTransforms* (ANTs), configured with Lanczos interpolation to minimize the smoothing effects of other kernels (Lanczos, 1964). Non-gridded (surface) resamplings were performed using *mri_vol2surf* (FreeSurfer). Many internal operations of *fMRIPrep* use *Nilearn* 0.9.1 (Abraham et al., 2014), mostly within the functional processing workflow.

The next preprocessing steps were performed using CONN (Whitfield-Gabrieli & Nieto-Castanon, 2012) release 22.a (Nieto-Castanon & Whitfield-Gabrieli, 2022) and SPM release 12.7771 (Penny et al., 2006). Functional data were smoothed using spatial convolution with a Gaussian kernel of 6 mm full width half maximum (FWHM) and functional data was masked with the built-in volumetric brain mask. In addition, functional data were denoised using a standard denoising pipeline (Nieto-Castanon, 2020) including the regression of potential confounding effects derived from the fMRIprep pipeline. The regression included artifacts from head motion with the Friston 24-parameter model encompassing six head motion parameters estimated from the current volume, the corresponding six motion parameters estimated from the immediately preceding volume, and twelve corresponding squared terms (Friston et al., 1996), physiological noise from white matter and CSF with the anatomical component correction (aCompCor) method (Behzadi et al., 2007), and scrubbing of volumes with a framewise displacement (FD) > 0.5 mm (Power et al., 2014). Additionally, the regression of global signal (GSR) was done as complementary analysis and results are reported both with and without GSR as the inclusion of GSR is still an ongoing debate in the (psychedelic) neuroimaging field (McCulloch et al., 2022). After these steps, the data was linearly detrended with 2 factors and bandpass-filtered (0.008–0.09 Hz) (Hallquist et al., 2013). From the number of noise terms included in this denoising strategy, the effective degrees of freedom of the BOLD signal after denoising were estimated to range from 0 to 77.3 (average 66.4) across all subjects (Nieto-Castanon, 2020).

### Suppl. Results

**Suppl. Table 1** Participant Characteristics of the analyzed sample (n=39).

| | DMT-harmine group | Placebo group | *p* | SMD |
|---|---|---|---|---|
| **n** | 19 | 20 | | |
| **Age (mean (SD))** | 41.37 (11.78) | 45.60 (8.38) | 0.202 | 0.414 |
| **Sex (%)** | | | | |
| male | 11 (57.9) | 11 (55.0) | 1.000 | 0.058 |
| female | 8 (42.1) | 9 (45.0) | | |
| **Hours of meditation practice (mean (SD))** | 2176.32 (1532.82) | 2720.00 (2182.37) | 0.376 | 0.288 |
| **Highest Education level (%)** | | | | |
| primary school degree | 2 (10.5) | 1 (5.0) | 0.72 | 0.380 |
| secondary school degree | 0 (0.0) | 1 (5.0) | | |
| high school degree | 1 (5.3) | 1 (5.0) | | |
| university degree | 16 (84.2) | 17 (85.0) | | |
| **Years of education (mean (SD))** | 17.95 (5.46) | 18.35 (5.34) | 0.817 | 0.075 |
| **Ethnicity (%)** | | | | |
| White | 18 (94.7) | 19 (95.0) | 1.000 | 0.012 |
| Hispanic | 1 (5.3) | 1 (5.0) | | |
| **Religious belief (%)** | | | | |
| Christian | 4 (21.1) | 3 (15.0) | 0.742 | 0.631 |
| Buddhist | 4 (21.1) | 3 (15.0) | | |
| Atheist | 0 (0.0) | 1 (5.0) | | |
| Pantheist/Animist | 0 (0.0) | 1 (5.0) | | |
| Spiritual, but not religious | 7 (36.8) | 9 (45.0) | | |
| None | 3 (15.8) | 3 (15.0) | | |
| Other | 1 (5.3) | 0 (0.0) | | |

Values are either given as mean (standard deviation) or as frequency (%) including standard mean difference (SMD) and *p*-value.

**Suppl. Table 2** Motion parameters per functional MRI scan. For each subject, the mean and maximum framewise displacement (FD, in mm) are reported, along with the number and percentage of volumes exceeding 0.5 mm FD relative to the preceding volume (total volumes = 240). Sub-12 exhibited excessive head motion in both scans and was excluded from all analyses (highlighted in orange).

| Participant | Condition | Timepoint | FD mean | FD max | # FD>0.5 mm | % FD>0.5mm |
|---|---|---|---|---|---|---|
| sub-01 | DMT-harmine | Pre | 0.22 | 0.52 | 1 | 0.4 |
| | | Post | 0.20 | 0.73 | 3 | 1.3 |
| sub-02 | DMT-harmine | Pre | 0.13 | 0.46 | - | - |
| | | Post | 0.21 | 0.72 | 4 | 1.7 |
| sub-03 | Placebo | Pre | 0.18 | 1.35 | 8 | 3.3 |
| | | Post | 0.25 | 1.84 | 22 | 9.2 |
| sub-04 | DMT-harmine | Pre | 0.18 | 0.80 | 4 | 1.7 |
| | | Post | 0.19 | 1.03 | 2 | 0.8 |
| sub-05 | Placebo | Pre | 0.25 | 0.60 | 8 | 3.3 |
| | | Post | 0.25 | 0.65 | 7 | 2.9 |
| sub-06 | DMT-harmine | Pre | 0.35 | 1.79 | 38 | 15.9 |
| | | Post | 0.27 | 1.08 | 9 | 3.8 |
| sub-07 | DMT-harmine | Pre | 0.20 | 0.47 | - | - |
| | | Post | 0.28 | 0.66 | 16 | 6.7 |
| sub-08 | Placebo | Pre | 0.18 | 0.46 | - | - |
| | | Post | 0.48 | 4.26 | 63 | 26.4 |
| sub-09 | Placebo | Pre | 0.17 | 0.38 | - | - |
| | | Post | 0.17 | 0.41 | - | - |
| sub-10 | DMT-harmine | Pre | 0.28 | 0.68 | 10 | 4.2 |
| | | Post | 0.30 | 0.66 | 10 | 4.2 |
| sub-11 | Placebo | Pre | 0.32 | 0.77 | 21 | 8.8 |
| | | Post | 0.25 | 0.76 | 7 | 2.9 |
| sub-12 | Placebo | Pre | 0.48 | 0.94 | 110 | 46.0 |
| | | Post | 0.48 | 0.87 | 107 | 44.8 |
| sub-13 | DMT-harmine | Pre | 0.19 | 0.51 | 1 | 0.4 |
| | | Post | 0.22 | 0.54 | 2 | 0.8 |
| sub-14 | DMT-harmine | Pre | 0.13 | 0.38 | - | - |
| | | Post | 0.12 | 0.35 | - | - |
| sub-15 | DMT-harmine | Pre | 0.28 | 0.92 | 8 | 3.3 |
| | | Post | 0.32 | 1.18 | 32 | 13.4 |
| sub-16 | Placebo | Pre | 0.25 | 0.61 | 8 | 3.3 |
| | | Post | 0.24 | 0.60 | 3 | 1.3 |
| sub-17 | DMT-harmine | Pre | 0.27 | 0.64 | 4 | 1.7 |
| | | Post | 0.25 | 0.55 | 3 | 1.3 |
| sub-18 | Placebo | Pre | 0.12 | 0.30 | - | - |
| | | Post | 0.13 | 0.40 | - | - |
| sub-19 | Placebo | Pre | 0.16 | 0.71 | 1 | 0.4 |
| | | Post | 0.20 | 0.48 | - | - |
| sub-20 | DMT-harmine | Pre | 0.19 | 0.59 | 1 | 0.4 |
| | | Post | 0.21 | 0.57 | 3 | 1.3 |

| | | | | | | |
|---|---|---|---|---|---|---|
| sub-21 | Placebo | Pre | 0.33 | 0.81 | 39 | 16.3 |
| | | Post | 0.27 | 0.83 | 15 | 6.3 |
| sub-22 | Placebo | Pre | 0.21 | 0.47 | - | - |
| | | Post | 0.26 | 0.68 | 5 | 2.1 |
| sub-23 | DMT-harmine | Pre | 0.22 | 0.90 | 8 | 3.3 |
| | | Post | 0.20 | 0.62 | 3 | 1.3 |
| sub-24 | DMT-harmine | Pre | 0.28 | 0.79 | 30 | 12.6 |
| | | Post | 0.25 | 0.73 | 6 | 2.5 |
| sub-25 | Placebo | Pre | 0.22 | 0.75 | 5 | 2.1 |
| | | Post | 0.15 | 0.39 | - | - |
| sub-26 | DMT-harmine | Pre | 0.35 | 1.28 | 54 | 22.6 |
| | | Post | 0.25 | 1.19 | 11 | 4.6 |
| sub-27 | DMT-harmine | Pre | 0.19 | 0.58 | 1 | 0.4 |
| | | Post | 0.16 | 1.10 | 4 | 1.7 |
| sub-28 | Placebo | Pre | 0.18 | 0.55 | 1 | 0.4 |
| | | Post | 0.15 | 0.57 | 2 | 0.8 |
| sub-29 | Placebo | Pre | 0.24 | 0.70 | 9 | 3.8 |
| | | Post | 0.26 | 0.59 | 7 | 2.9 |
| sub-30 | DMT-harmine | Pre | 0.27 | 0.61 | 6 | 2.5 |
| | | Post | 0.22 | 0.50 | - | - |
| sub-31 | Placebo | Pre | 0.20 | 0.53 | 1 | 0.4 |
| | | Post | 0.18 | 0.56 | 1 | 0.4 |
| sub-32 | DMT-harmine | Pre | 0.14 | 0.45 | - | - |
| | | Post | 0.21 | 0.53 | 2 | 0.8 |
| sub-33 | Placebo | Pre | 0.30 | 1.90 | 32 | 13.4 |
| | | Post | 0.31 | 0.82 | 26 | 10.9 |
| sub-34 | Placebo | Pre | 0.30 | 4.75 | 11 | 4.6 |
| | | Post | 0.20 | 0.90 | 3 | 1.3 |
| sub-35 | Placebo | Pre | 0.21 | 1.73 | 2 | 0.8 |
| | | Post | 0.17 | 0.69 | 5 | 2.1 |
| sub-36 | Placebo | Pre | 0.23 | 0.58 | 2 | 0.8 |
| | | Post | 0.31 | 0.79 | 19 | 7.9 |
| sub-37 | DMT-harmine | Pre | 0.17 | 0.42 | - | - |
| | | Post | 0.21 | 0.48 | - | - |
| sub-38 | DMT-harmine | Pre | 0.22 | 2.25 | 7 | 2.9 |
| | | Post | 0.19 | 1.01 | 1 | 0.4 |
| sub-39 | Placebo | Pre | 0.18 | 0.49 | - | - |
| | | Post | 0.16 | 0.38 | - | - |
| sub-40 | DMT-harmine | Pre | 0.15 | 0.40 | - | - |
| | | Post | 0.14 | 0.42 | - | - |

**Suppl. Table 3** Mean motion parameters per condition and timepoint. For each group, the mean and maximum framewise displacement (FD, in mm) are reported, along with the number and percentage of volumes exceeding 0.5 mm FD relative to the preceding volume (total volumes = 240 x number of participants in group).

| Condition | Timepoint | Group size | FD mean | FD max | # FD>0.5 mm | % FD>0.5mm |
|---|---|---|---|---|---|---|
| DMT-harmine | Pre | 19 | 0.24 | 4.75 | 258 | 5.4 |
| | Post | 19 | 0.25 | 4.26 | 292 | 6.1 |
| Placebo | Pre | 20 | 0.22 | 2.25 | 173 | 3.6 |
| | Post | 20 | 0.22 | 1.19 | 111 | 2.3 |

**Suppl. Table 4** Results of repeated-measures ANOVA on mean framewise displacement, examining the main effects of Condition and Timepoint, and their interaction. Shown are the Sum of Squares (*SS*), degrees of freedom (*df*), Mean Squares (*MS*), *F*-values, uncorrected p-values (*p*), and partial eta squared ($\eta^2$) as a measure of effect size.

| Factor | *SS* | *df* | *MS* | *F* | *p* | $\eta^2$ |
|---|---|---|---|---|---|---|
| **Condition** | 0.00109 | (1,37) | 0.00109 | 0.181 | 0.673 | 0.005 |
| **Timepoint** | 0.0003 | (1,37) | 0.0003 | 0.127 | 0.724 | 0.003 |
| **Interaction** | 0.00052 | (1,37) | 0.00052 | 0.220 | 0.642 | 0.006 |

**Suppl. Table 5** Smoothness estimates derived from first-level analysis (with *CONN*) for the network analysis shown in Figure 3 in the main manuscript. Estimates are given in mm in x,y,z direction for each contrast and network that is shown to have significant clusters in Figure 3. Abbreviations: VIS – visual network, DAN – dorsal attention network, SAL – salience network, FPN – frontoparietal network.

| Contrast | RSN | x | y | z |
|---|---|---|---|---|
| **Synergy** | VIS | 4.31 | 4.29 | 4.01 |
| | SAL | 4.24 | 4.22 | 3.95 |
| **Group differences post retreat** | SAL | 4.45 | 4.47 | 4.18 |
| **Meditation** | VIS | 4.31 | 4.29 | 4.01 |
| | DAN | 4.27 | 4.24 | 3.97 |
| | FPN | 4.26 | 4.26 | 3.97 |
| **Psychedelic-augmented meditation** | SAL | 4.27 | 4.24 | 3.97 |

**Suppl. Table 6** Within-network FC values for each condition.

**Without GSR**

| RSN | DMT-harmine Pre | Placebo Pre | DMT-harmine Post | Placebo Post |
|---|---|---|---|---|
| **VIS *** | 0.442 (0.086) | 0.438 (0.092) | 0.516 (0.124) | 0.477 (0.096) |
| **ASM** | 0.440 (0.094) | 0.437 (0.064) | 0.473 (0.125) | 0.448 (0.083) |
| **DAN** | 0.379 (0.063) | 0.383 (0.047) | 0.386 (0.105) | 0.358 (0.057) |
| **SAL** | 0.446 (0.078) | 0.452 (0.060) | 0.452 (0.131) | 0.418 (0.068) |
| **FPN** | 0.350 (0.060) | 0.349 (0.058) | 0.351 (0.100) | 0.334 (0.059) |
| **DMN** | 0.369 (0.051) | 0.394 (0.064) | 0.373 (0.107) | 0.372 (0.061) |

**With GSR**

| RSN | DMT-harmine Pre | Placebo Pre | DMT-harmine Post | Placebo Post |
|---|---|---|---|---|
| **VIS *** | 0.316 (0.074) | 0.314 (0.071) | 0.360 (0.084) | 0.368 (0.082) |
| **ASM** | 0.350 (0.070) | 0.337 (0.051) | 0.352 (0.059) | 0.344 (0.068) |
| **DAN** | 0.300 (0.045) | 0.289 (0.042) | 0.277 (0.035) | 0.273 (0.052) |
| **SAL** | 0.381 (0.047) | 0.385 (0.064) | 0.381 (0.080) | 0.363 (0.063) |
| **FPN** | 0.280 (0.054) | 0.269 (0.046) | 0.276 (0.058) | 0.281 (0.056) |
| **DMN** | 0.295 (0.038) | 0.316 (0.052) | 0.299 (0.048) | 0.303 (0.060) |

Within-network FC values are shown as mean and standard deviation. **Two-way repeated measures ANOVAs with interaction, independent samples, and paired *t*-tests** were computed on *CONN*-derived FC values. * - depicts significant differences between conditions for uncorrected *p*-values. Abbreviations: VIS – visual network, ASM – auditory-sensorimotor network, DAN – dorsal attention network, SAL – salience network, FPN – frontoparietal network, DMN – default mode network.

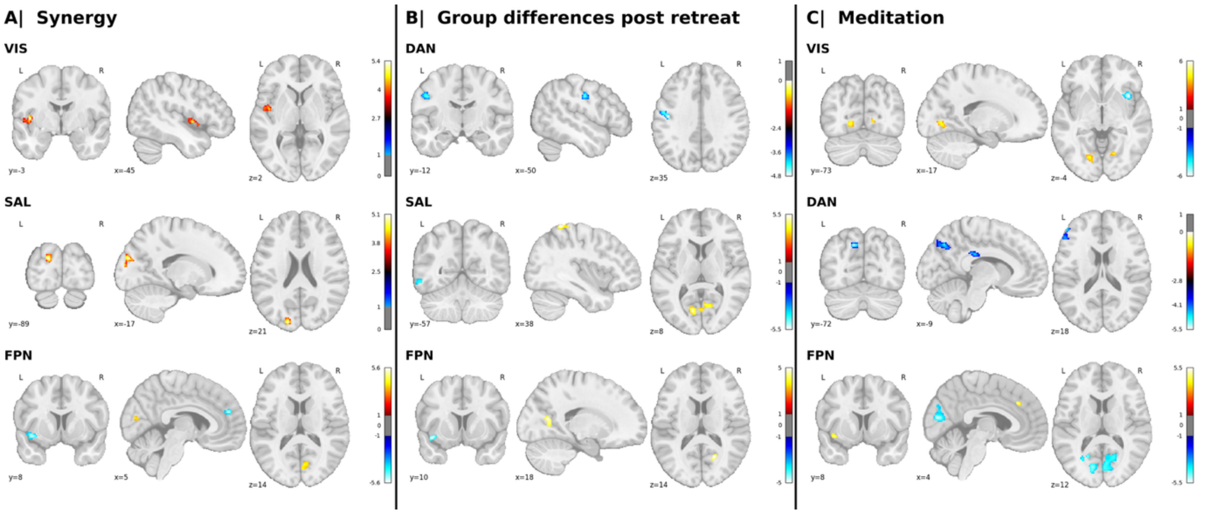


**Suppl. Figure 1 Changes in network connectivity between groups and timepoints with global signal regression.** Voxel-wise statistical parametric maps that represent changes in connectivity between one RSN (i.e., the seed) and the whole brain are shown. The allocation of each significant cluster to its corresponding Yeo network is indicated in parentheses. The panels correspond to different contrasts between drug condition (i.e. DMT-harmine vs placebo) and timepoint (i.e., pre vs post meditation retreat). **A| Synergy** (DMT-harmine > placebo: Post > pre): The two-way repeated measures ANOVA model shows increased FC between VIS and SAL (with both VIS and SAL as seed) and between FPN and VIS and decreased FC between FPN and SAL. **B| Group differences post retreat** (DMT-harmine > Placebo: Post): After the retreat, an ANCOVA showed decreased FC between DAN and ASM, and increased FC between SAL and VIS and between FPN and VIS. **C| Meditation** (Placebo: Post > Pre: ANCOVA indicated decreased FC within the placebo group after the retreat between VIS and SAL, DAN and DMN and FPN and VIS, as well as increased FC within VIS and FPN, and between FPN and DMN. The analyses were computed in CONN ($p<0.001$, cluster-level FDR-adjusted $q_{FDR}<.05$); x, y, and z indicate Montreal Neurological Institute (MNI) coordinates, warm colors represent increased connectivity and cold colors decreased connectivity for the given contrast.

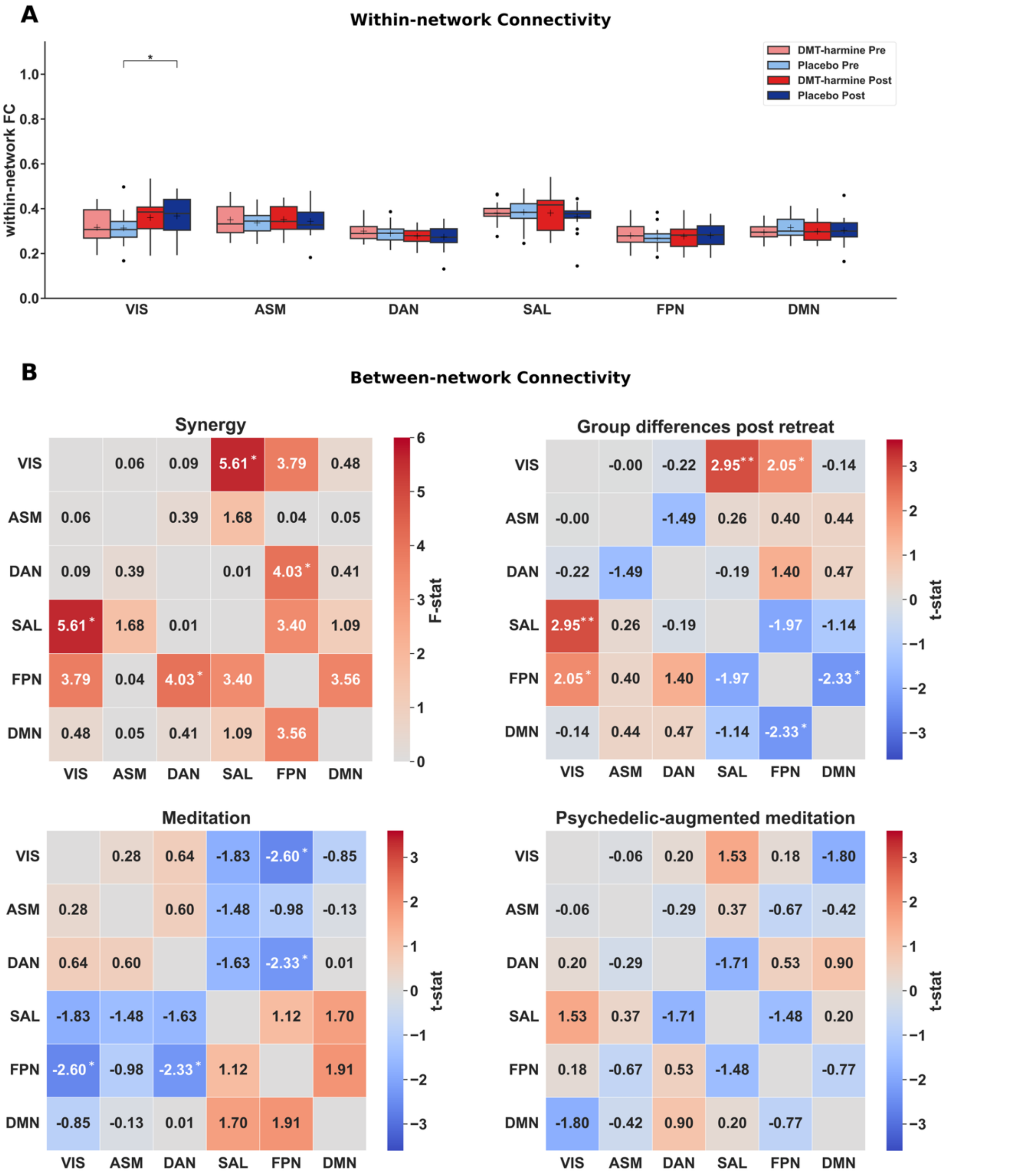


**Suppl. Figure 2 Within-and between-network FC changes after verum (DMT + harmine) or placebo with global signal regression. A|** No significant results were observed for the within-network FC analysis following multiple correction; however, the following trends were observed at the uncorrected level. Within-network FC analysis showed a significant main effect of time in VIS in the **Synergy** contrast (*F*(1,74)=7.77, $p_{uncorrected}$=.007*)*. A significant increase in within-VIS FC was observed following the retreat in the *placebo* group in the **Meditation** contrast (*t*=*2.20*, $p_{uncorrected}$=.0*31*). No other significant within-network effects were detected for other contrasts (i.e., **Group differences post retreat** and **Psychedelic-augmented meditation**). **C|** Similarly, for between-network FC analysis no results survived correction for multiple testing. However, uncorrected trend-level effects were observed. Repeated-measures ANOVA (**Synergy** contrast, top left matrix) showed significant interactions between VIS-SAL (*F*(1,74)=5.61, $p_{uncorrected}$=.02*1*) and DAN-FPN (*F*(1,74)=4.03, $p_{uncorrected}$=.*048*) for $p_{uncorrected}$<.05. In the **Group differences post retreat** contrast, increased FC was observed between VIS-SAL (*t*=*2.95*, $p_{uncorrected}$=.*004*) and VIS-FPN (*t*=*2.05*, $p_{uncorrected}$=.*043*) *as well as decreased FC between FPN-DMN (t*=2.33, $p_{uncorrected}$=.022) in the DMT-harmine group. The **Meditation** contrast showed decreased FC between VIS-FPN (*t*=2.60, $p_{uncorrected}$=.072) and DAN-FPN (*t*=2.33, $p_{uncorrected}$=.023) after the retreat. Asterisks denote significant differences for differences in t-tests for the given contrast without adjustment for multiple comparisons (*: $p_{uncorrected}$<.05). Abbreviations: VIS – visual network, ASM – auditory-sensorimotor network, DAN – dorsal attention network, SAL – salience network, FPN – frontoparietal network, DMN – default mode network.

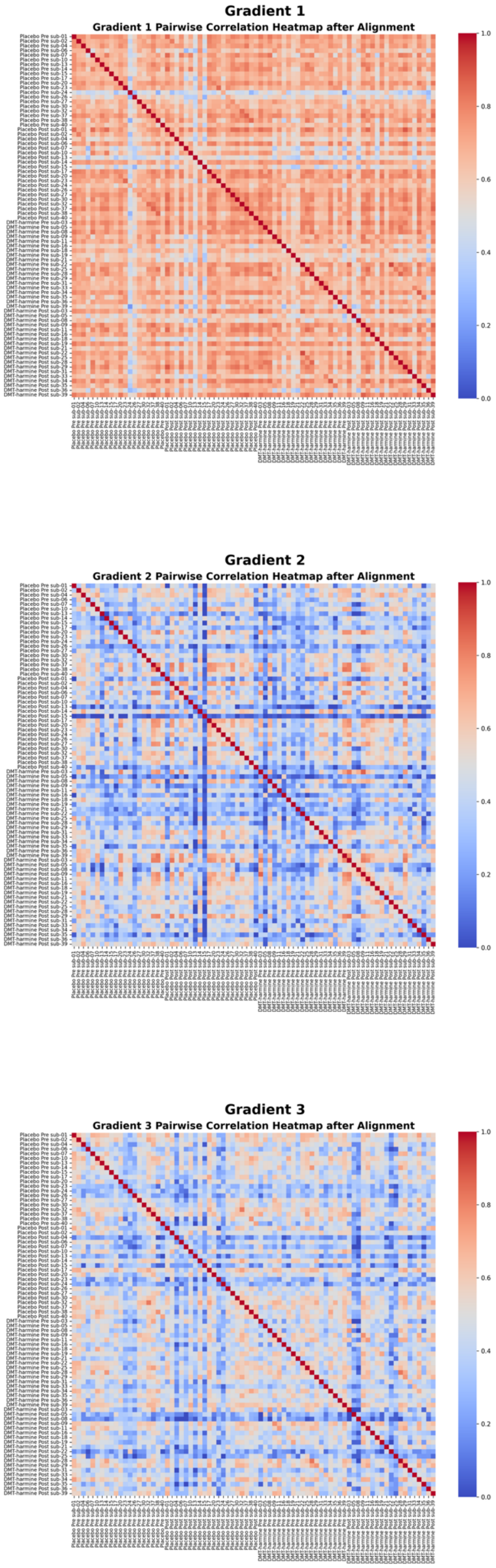


**Suppl. Figure 3 Pairwise correlation heatmaps of aligned gradient components across subjects.** This figure displays the Pearson correlation matrices for the first three gradients after Procrustes alignment for each scan. Each heatmap illustrates the similarity in gradient profiles between all pairs of subjects, with warmer colors (red) indicating higher correlation and cooler colors (blue) indicating lower correlation. This visualization highlights the consistency and individual variability of gradient components across the sample, showing that the alignment correlates strongest within the first cortical gradient (top) and to a lesser extent within the second (middle) and third (bottom) cortical gradients. This heatmap corresponds to the data shown in the manuscript in **Figure 4**, i.e. without inclusion of global signal regression.

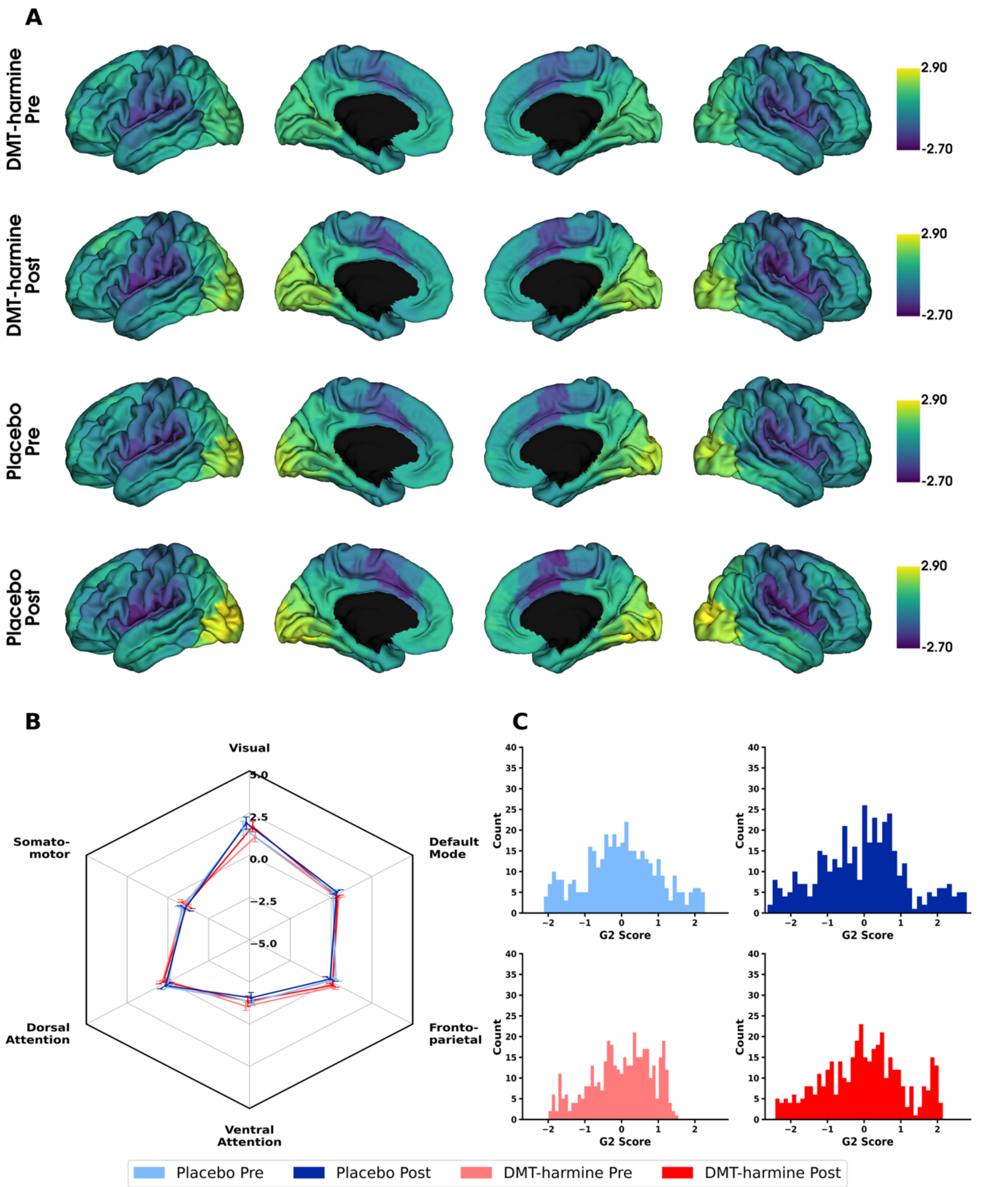


**Suppl. Figure 4 Cortical second gradients before and after retreat for DMT-harmine and placebo group. A|** Parcellation-wise mean cortical second gradients for *DMT-harmine Pre, DMT-harmine Post, Placebo Pre and Placebo Post* conditions that represent the axis from visual to insular cortex. The shown gradient scores represent arbitrary units, with negative values defining the lower end of the gradient axis (i.e., visual areas) and higher (positive) numbers representing the higher end of the axis (i.e., insular cortices). **B|** Network-wise mean cortical gradients. The same data is presented network-based, wherein each brain parcel was allocated to the corresponding Yeo network. Mean and SEM are presented per network. **C|** Histograms showing the distribution of gradient values for each condition for each brain parcellation. Statistical differences were tested with a two-way repeated measures ANOVA (for within- and between-group effects and their interactions). No significant differences were observed for any contrast, neither with parcellation- nor network-wise contrasts.

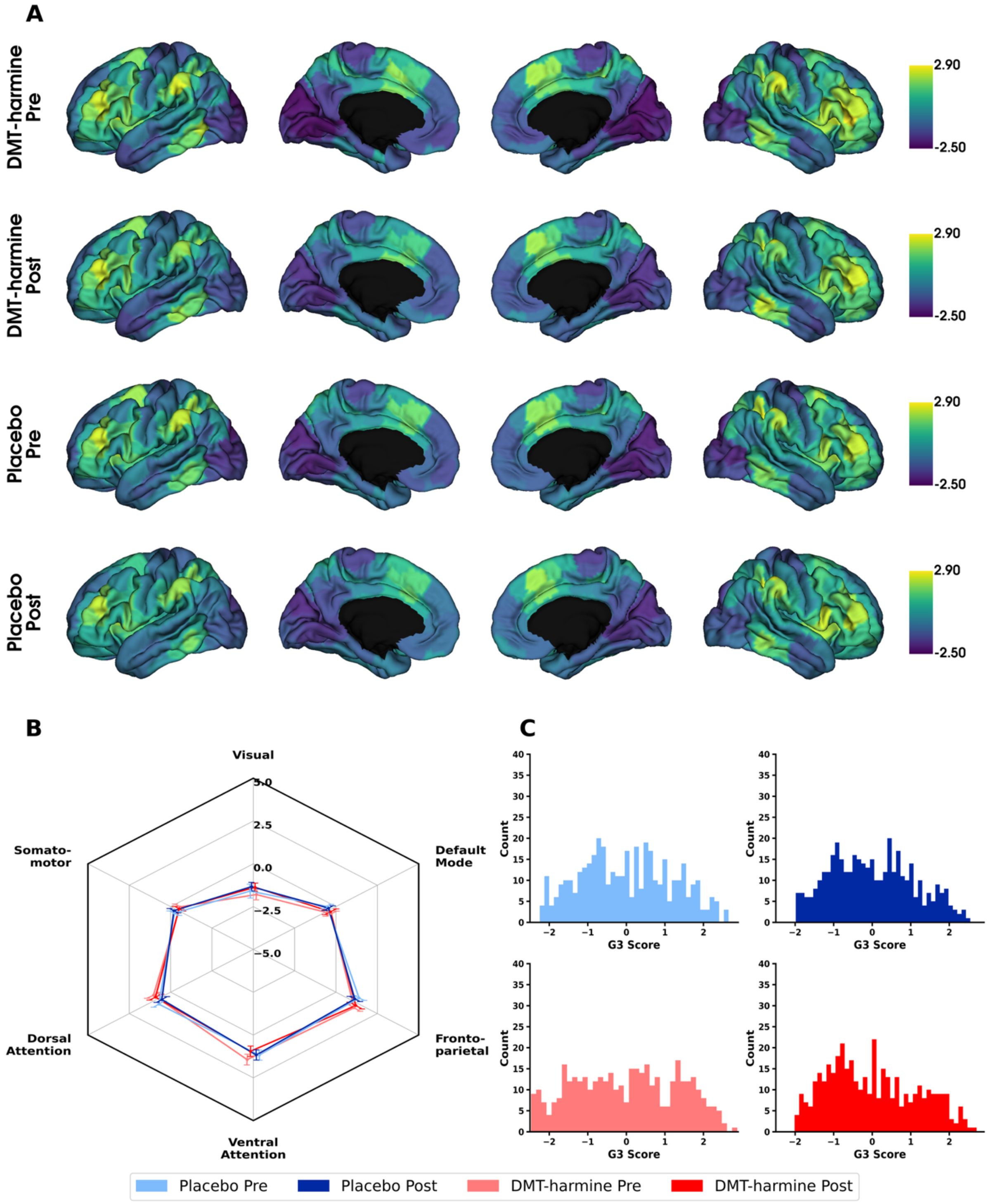


**Suppl. Figure 5 Cortical third gradients before and after retreat for DMT-harmine and placebo group. A|** Parcellation-wise mean cortical second gradients for *DMT-harmine Pre, DMT-harmine Post, Placebo Pre and Placebo Post* conditions that represent the axis from visual to frontoparietal network. The shown gradient scores represent arbitrary units, with negative values defining the lower end of the gradient axis (i.e., visual areas) and higher (positive) numbers representing the higher end of the axis (i.e., regions covered by the frontoparietal network). **B|** Network-wise mean cortical gradients. The same data is presented network-based, wherein each brain parcel was allocated to the corresponding Yeo network. Mean and SEM are presented per network. **C|** Histograms showing the distribution of gradient values for each condition for each brain parcellation. Statistical differences were tested with a two-way repeated measures ANOVA (for within- and between-group effects and their interactions). No significant differences were observed for any contrast, neither with parcellation- nor network-wise contrasts.

**Suppl. Table 7** Results from two-way ANOVA on 3D within-network dispersion within canonical functional networks. Within-network dispersion quantifies the sum squared Euclidean distance of parcels to their respective network centroids in individualized gradient space. The analysis tested for main effects of Drug condition (within-subject factor: DMT + harmine, Placebo), Timepoint (between-subject factor: Pre, Post), and their interaction. Reported are *F*-values and FDR-corrected *p*-values (*qFDR*) for each effect across six Yeo networks (LIM excluded).

| | **Drug condition** | | **Timepoint** | | **Interaction** | |
|---|---|---|---|---|---|---|
| **network** | ***F*** | $q_{FDR}$ | ***F*** | $q_{FDR}$ | ***F*** | $q_{FDR}$ |
| **VIS** | 1.569 | 0.549 | 1.274 | 0.529 | 0.059 | 0.996 |
| **ASM** | 0.021 | 0.886 | 0.641 | 0.529 | 0.000 | 0.996 |
| **DAN** | 0.038 | 0.886 | 0.991 | 0.529 | 0.003 | 0.996 |
| **SAL** | 1.211 | 0.549 | 4.209 | 0.262 | 1.113 | 0.996 |
| **FPN** | 1.439 | 0.549 | 0.527 | 0.529 | 0.772 | 0.996 |
| **DMN** | 0.524 | 0.707 | 0.401 | 0.529 | 0.071 | 0.996 |

**Suppl. Table 8** Results from two-way ANOVA on 3D between-network dispersion within canonical functional networks. Between-network dispersion quantifies the Euclidean distance between network centroids in individualized gradient space. The analysis tested for main effects of Drug condition (within-subject factor: DMT + harmine, Placebo), Timepoint (between-subject factor: Pre, Post), and their interaction. Reported are *F*-values and FDR-corrected *p*-values (*qFDR*) for each effect across six Yeo networks (LIM excluded).

| | **Drug condition** | | **Timepoint** | | **Interaction** | |
|---|---|---|---|---|---|---|
| **Network pair** | ***F*** | $q_{FDR}$ | ***F*** | $q_{FDR}$ | ***F*** | $q_{FDR}$ |
| **VIS-ASM** | 0.738 | 0.934 | 0.938 | 0.702 | 0.006 | 0.938 |
| **VIS-DAN** | 0.229 | 0.934 | 0.001 | 0.981 | 0.070 | 0.938 |
| **VIS-SAL** | 0.100 | 0.934 | 0.405 | 0.777 | 0.080 | 0.938 |
| **VIS-FPN** | 0.002 | 0.967 | 1.197 | 0.702 | 0.057 | 0.938 |
| **VIS-DMN** | 0.116 | 0.934 | 0.941 | 0.702 | 0.055 | 0.938 |
| **ASM-DAN** | 1.417 | 0.934 | 0.245 | 0.777 | 0.015 | 0.938 |
| **ASM-SAL** | 0.956 | 0.934 | 0.026 | 0.981 | 0.672 | 0.938 |
| **ASM-FPN** | 0.067 | 0.934 | 1.421 | 0.702 | 0.363 | 0.938 |
| **ASM-DMN** | 0.045 | 0.934 | 0.798 | 0.702 | 0.634 | 0.938 |
| **DAN-SAL** | 1.254 | 0.934 | 6.394 | 0.204 | 0.234 | 0.938 |
| **DAN-FPN** | 0.857 | 0.934 | 1.698 | 0.702 | 0.931 | 0.938 |
| **DAN-DMN** | 0.839 | 0.934 | 0.304 | 0.777 | 0.511 | 0.938 |
| **FPN-SAL** | 0.211 | 0.934 | 2.250 | 0.702 | 0.011 | 0.938 |
| **FPN-DMN** | 0.290 | 0.934 | 0.333 | 0.777 | 0.085 | 0.938 |

**Suppl. Table 9** Ordinary least squares regression results for questionnaires EBI, MEQ, NADA-S, PIS, TMS.

| Questionnaire | Predictor | Coefficient | 95% CI | $q_{FDR}$ | $R^2$ |
|---|---|---|---|---|---|
| **MEQ** | Intercept | 1.0369 | 0.4549 – 1.6188 | 0.002 | 0.47 |
| | Group | -0.5566 | -0.8654 – -0.2478 | 0.002 | |
| | Total Score | -0.0102 | -0.0191 – -0.0012 | 0.068 | |
| | Total Score:Group | 0.0065 | 0.0014 – 0.0117 | **0.036*** | |
| **EBI** | Intercept | 0.821 | 0.3734 – 1.2686 | 0.002 | 0.434 |
| | Group | -0.4235 | -0.6620 – -0.1850 | 0.002 | |
| | Total Score | -0.0082 | -0.0171 – 0.0007 | 0.088 | |
| | Total Score:Group | 0.005 | 0.0000 – 0.0100 | 0.062 | |
| **NADA-S** | Intercept | 0.9814 | 0.4144 – 1.5484 | 0.002 | 0.441 |
| | Group | -0.5123 | -0.8137 – -0.2109 | 0.002 | |
| | Total Score | -0.008 | -0.0160 – -0.0001 | 0.079 | |
| | Total Score:Group | 0.0048 | 0.0003 – 0.0094 | 0.060 | |
| **PIS** | Intercept | 0.4914 | 0.1419 – 0.8409 | 0.007 | 0.365 |
| | Group | -0.2367 | -0.4363 – -0.0370 | 0.022 | |
| | Total Score | -0.0008 | -0.0081 – 0.0066 | 0.836 | |
| | Total Score:Group | 0.0004 | -0.0038 – 0.0047 | 0.835 | |
| **TMS** | Intercept | 1.5359 | 0.6070 – 2.4648 | 0.002 | 0.468 |
| | Group | -0.8928 | -1.4303 – -0.3553 | 0.002 | |
| | Total Score | -0.0142 | -0.0264 – -0.0021 | 0.068 | |
| | Total Score:Group | 0.0089 | 0.0019 – 0.0159 | **0.036*** | |

Averaged timeseries derived from the fMRI cluster from the contrast *DMT-harmine > Placebo: Post* showing increased FC between salience and visual network were used as the dependent variable. Group, Total Score (%), and Total Score:Group interaction were entered as predictors. *p*-values were corrected for the total number of questionnaires modeled (5) with FDR. The corresponding $q_{FDR}$ values are shown. * - indicates significant ($q_{FDR}$<.05) Total Score:Group interactions. Abbreviations: MEQ – Mystical Experience Questionnaire, EBI – Emotional Breakthrough Inventory, NADA-S – Nondual Awareness Dimensional Assessment, PIS – Psychological Insight Scale, TMS – Toronto Mindfulness Scale.

**Suppl. Table 10** Results from exploratory post-hoc correlational analyses for questionnaires EBI, MEQ, NADA-S, PIS, TMS.

| Questionnaire | Type | Group | Coefficient | $p_{uncorr}$ |
|---|---|---|---|---|
| **MEQ** | Pearson | DMT-harmine | -0.34 | 0.16 |
| | | Placebo | 0.52 | **0.020*** |
| **EBI** | Spearman | DMT-harmine | -0.15 | 0.542 |
| | | Placebo | 0.46 | **0.041*** |
| **NADA-S** | Pearson | DMT-harmine | -0.34 | 0.156 |
| | | Placebo | 0.36 | 0.118 |
| **PIS** | Pearson | DMT-harmine | -0.04 | 0.874 |
| | | Placebo | 0.03 | 0.905 |
| **TMS** | Spearman | DMT-harmine | -0.35 | 0.142 |
| | | Placebo | 0.27 | 0.247 |

Averaged timeseries derived from the fMRI cluster from the contrast *DMT-harmine > Placebo: Post* showing increased FC between salience and visual network were correlated with the Total Score (%) of questionnaires MEQ, EBI, NADA-S, PIS, and TMS. Correlations were done separately per group. Exploratory *p*-values were not corrected for multiple testing. * - indicates significant (*p*<.05) correlations. Abbreviations: MEQ – Mystical Experience Questionnaire, EBI – Emotional Breakthrough Inventory, NADA-S – Nondual Awareness Dimensional Assessment, PIS – Psychological Insight Scale, TMS – Toronto Mindfulness Scale.

**Suppl. Table 11** Ordinary least squares regression results for differences in questionnaire scores from baseline to post drug intervention at Day 2 for EBI, MEQ, NADA-S, PIS, TMS.

| Questionnaire | Predictor | Coefficient | 95% CI | $q_{FDR}$ | $R^2$ |
|---|---|---|---|---|---|
| **Δ MEQ** | Intercept | 0.6136 | 0.3267 – 0.9004 | 0.000 | 0.399 |
| | Group | -0.2984 | -0.4539 – -0.1429 | 0.001 | |
| | Total Score | -0.0058 | -0.0140 – 0.0025 | 0.278 | |
| | Total Score:Group | 0.0035 | -0.0018 – 0.0088 | 0.314 | |
| **Δ EBI** | Intercept | 0.7423 | 0.3777 – 1.1069 | 0.000 | 0.427 |
| | Group | -0.3733 | -0.5684 – -0.1781 | 0.001 | |
| | Total Score | -0.0083 | -0.0170 – 0.0004 | 0.155 | |
| | Total Score:Group | 0.0052 | -0.0002 – 0.0107 | 0.150 | |
| **Δ NADA-S** | Intercept | 0.6895 | 0.4653 – 0.9136 | 0.000 | 0.475 |
| | Group | -0.3360 | -0.4623 – -0.2097 | 0.000 | |
| | Total Score | -0.0063 | -0.0112 – -0.0014 | 0.067 | |
| | Total Score:Group | 0.0034 | 0.0003 – 0.0064 | 0.150 | |
| **Δ PIS** | Intercept | 0.4201 | 0.2038 – 0.6364 | 0.000 | 0.376 |
| | Group | -0.1949 | -0.3200 – -0.0698 | 0.003 | |
| | Total Score | 0.0020 | -0.0041 – 0.0081 | 0.508 | |
| | Total Score:Group | -0.0014 | -0.0052 – 0.0023 | 0.516 | |
| **Δ TMS** | Intercept | 0.5366 | 0.2707 – 0.8026 | 0.000 | 0.374 |
| | Group | -0.2599 | -0.4161 – -0.1038 | 0.002 | |
| | Total Score | -0.0043 | -0.0164 – 0.0078 | 0.508 | |
| | Total Score:Group | 0.0024 | -0.0049 – 0.0096 | 0.516 | |

Averaged timeseries derived from the fMRI cluster from the contrast *DMT-harmine > Placebo: Post* showing increased FC between salience and visual network were used as the dependent variable. Group, ΔTotal Score (%) from Baseline (either Day 0 or Day 1 depending on questionnaire) to Day 2, and ΔTotal Score:Group interaction were entered as predictors. *p*-values were corrected for the total number of questionnaires modeled (5) with FDR. The corresponding $q_{FDR}$ values are shown. * - indicates significant ($q_{FDR}$<.05) Total Score:Group interactions. Abbreviations: MEQ – Mystical Experience Questionnaire, EBI – Emotional Breakthrough Inventory, NADA-S – Nondual Awareness Dimensional Assessment, PIS – Psychological Insight Scale, TMS – Toronto Mindfulness Scale.

**Suppl. Table 12** Results from exploratory post-hoc correlational analyses for differences in questionnaire scores from baseline to post drug intervention at Day 2 for EBI, MEQ, NADA-S, PIS, TMS.

| Questionnaire | Type | Group | Coefficient | $p_{uncorr}$ |
|---|---|---|---|---|
| **Δ MEQ** | Pearson | DMT-harmine | -0.31 | 0.190 |
| | | Placebo | 0.31 | 0.177 |
| **Δ EBI** | Spearman | DMT-harmine | -0.15 | 0.547 |
| | | Placebo | 0.05 | 0.835 |
| **Δ NADA-S** | Pearson | DMT-harmine | 0.09 | 0.712 |
| | | Placebo | -0.19 | 0.427 |
| **Δ PIS** | Pearson | DMT-harmine | -0.25 | 0.293 |
| | | Placebo | 0.18 | 0.437 |
| **Δ TMS** | Spearman | DMT-harmine | -0.51 | **0.025*** |
| | | Placebo | 0.11 | 0.651 |

Averaged timeseries derived from the fMRI cluster from the contrast *DMT-harmine > Placebo: Post* showing increased FC between salience and visual network were correlated with ΔTotal Score (%) of the questionnaires MEQ, EBI, NADA-S, PIS, and TMS. Correlations were done separately per group. Exploratory *p*-values were not corrected for multiple testing. * - indicates significant ($p<.05$) correlations. Abbreviations: MEQ – Mystical Experience Questionnaire, EBI – Emotional Breakthrough Inventory, NADA-S – Nondual Awareness Dimensional Assessment, PIS – Psychological Insight Scale, TMS – Toronto Mindfulness Scale.

### References


Abraham, A., Pedregosa, F., Eickenberg, M., Gervais, P., Mueller, A., Kossaifi, J., Gramfort, A., Thirion, B., & Varoquaux, G. (2014). Machine learning for neuroimaging with scikit-learn. *Frontiers in Neuroinformatics*, *8*(FEB), 71792. https://doi.org/10.3389/FNINF.2014.00014/BIBTEX

Avants, B. B., Epstein, C. L., Grossman, M., & Gee, J. C. (2008). Symmetric diffeomorphic image registration with cross-correlation: Evaluating automated labeling of elderly and neurodegenerative brain. *Medical Image Analysis*, *12*(1), 26–41. https://doi.org/10.1016/J.MEDIA.2007.06.004

Behzadi, Y., Restom, K., Liau, J., & Liu, T. T. (2007). A component based noise correction method (CompCor) for BOLD and perfusion based fMRI. *NeuroImage*, *37*(1), 90–101. https://doi.org/10.1016/J.NEUROIMAGE.2007.04.042

Ciric, R., Thompson, W. H., Lorenz, R., Goncalves, M., MacNicol, E. E., Markiewicz, C. J., Halchenko, Y. O., Ghosh, S. S., Gorgolewski, K. J., Poldrack, R. A., & Esteban, O. (2022). TemplateFlow: FAIR-sharing of multi-scale, multi-species brain models. *Nature Methods 2022 19:12*, *19*(12), 1568–1571. https://doi.org/10.1038/s41592-022-01681-2

Cox, R. W., & Hyde, J. S. (1997). Software tools for analysis and visualization of fMRI data. *NMR in Biomedicine*, *10*(4–5), 171–178. https://doi.org/https://doi.org/10.1002/(SICI)1099-1492(199706/08)10:4/5<171::AID-NBM453>3.0.CO;2-L

Dale, A. M., Fischl, B., & Sereno, M. I. (1999). Cortical Surface-Based Analysis: I. Segmentation and Surface Reconstruction. *NeuroImage*, *9*(2), 179–194. https://doi.org/10.1006/NIMG.1998.0395

Esteban, O., Markiewicz, C. J., Blair, R. W., Moodie, C. A., Isik, A. I., Erramuzpe, A., Kent, J. D., Goncalves, M., DuPre, E., Snyder, M., Oya, H., Ghosh, S. S., Wright, J., Durnez, J., Poldrack, R. A., & Gorgolewski, K. J. (2019). fMRIPrep: a robust preprocessing pipeline for functional MRI. *Nature Methods*, *16*(1), 111–116. https://doi.org/10.1038/s41592-018-0235-4

Esteban, O., Markiewicz, C. J., Burns, C., Goncalves, M., Jarecka, D., Ziegler, E., Berleant, S., Ellis, D. G., Pinsard, B., Madison, C., Waskom, M., Notter, M. P., Clark, D., Manhães-Savio, A., Clark, D., Jordan, K., Dayan, M., Halchenko, Y. O., Loney, F., … Ghosh, S. (2022). *nipy/nipype: 1.8.3*. Zenodo. https://doi.org/10.5281/zenodo.6834519

Esteban, O., Markiewicz, C. J., Goncalves, M., Provins, C., Kent, J. D., DuPre, E., Salo, T., Ciric, R., Pinsard, B., Blair, R. W., Poldrack, R. A., & Gorgolewski, K. J. (2023). *fMRIPrep 23.0.2.* Zenodo. https://doi.org/10.5281/zenodo.7863421

Fonov, V., Evans, A., McKinstry, R., Almli, C., & Collins, D. (2009). Unbiased nonlinear average age-appropriate brain templates from birth to adulthood. *NeuroImage*, *47*, S102. https://doi.org/10.1016/S1053-8119(09)70884-5

Friston, K. J., Williams, S., Howard, R., Frackowiak, R. S. J., & Turner, R. (1996). Movement-Related effects in fMRI time-series. *Magnetic Resonance in Medicine*, *35*(3), 346–355. https://doi.org/10.1002/mrm.1910350312

Gorgolewski, K., Burns, C. D., Madison, C., Clark, D., Halchenko, Y. O., Waskom, M. L., & Ghosh, S. S. (2011). Nipype: A flexible, lightweight and extensible neuroimaging data processing framework in Python. *Frontiers in Neuroinformatics*, *5*, 12318. https://doi.org/10.3389/FNINF.2011.00013/ABSTRACT

Greve, D. N., & Fischl, B. (2009). Accurate and robust brain image alignment using boundary-based registration. *NeuroImage*, *48*(1), 63–72. https://doi.org/10.1016/J.NEUROIMAGE.2009.06.060

Hallquist, M. N., Hwang, K., & Luna, B. (2013). The nuisance of nuisance regression: Spectral misspecification in a common approach to resting-state fMRI preprocessing reintroduces noise and obscures functional connectivity. *NeuroImage*, *82*, 208–225. https://doi.org/10.1016/j.neuroimage.2013.05.116

Jenkinson, M., Bannister, P., Brady, M., & Smith, S. (2002). Improved Optimization for the Robust and Accurate Linear Registration and Motion Correction of Brain Images. *NeuroImage*, *17*(2), 825–841. https://doi.org/10.1006/NIMG.2002.1132

Klein, A., Ghosh, S. S., Bao, F. S., Giard, J., Häme, Y., Stavsky, E., Lee, N., Rossa, B., Reuter, M., Chaibub Neto, E., & Keshavan, A. (2017). Mindboggling morphometry of human brains. *PLOS Computational Biology*, *13*(2), e1005350. https://doi.org/10.1371/JOURNAL.PCBI.1005350

Lanczos, C. (1964). Evaluation of Noisy Data. *Journal of the Society for Industrial and Applied Mathematics Series B Numerical Analysis*, *1*(1), 76–85. https://doi.org/10.1137/0701007

McCulloch, D. E.-W., Knudsen, G. M., Barrett, F. S., Doss, M. K., Carhart-Harris, R. L., Rosas, F. E., Deco, G., Kringelbach, M. L., Preller, K. H., Ramaekers, J. G., Mason, N. L., Müller, F., & Fisher, P. M. (2022). Psychedelic resting-state neuroimaging: A review and perspective on balancing replication and novel analyses. *Neuroscience & Biobehavioral Reviews*, *138*, 104689. https://doi.org/10.1016/j.neubiorev.2022.104689

Nieto-Castanon, A. (2020). *Handbook of functional connectivity Magnetic Resonance Imaging methods in CONN*. Hilbert Press. https://doi.org/10.56441/hilbertpress.2207.6598

Nieto-Castanon, A., & Whitfield-Gabrieli, S. (2022). *CONN functional connectivity toolbox: RRID SCR_009550, release 22*. Hilbert Press. https://doi.org/10.56441/hilbertpress.2246.5840

Penny, W. D., Friston, K. J., Ashburner, J. T., Kiebel, S. J., & Nichols, T. E. (Eds.). (2006). *Statistical parametric mapping: the analysis of functional brain images*. Elsevier. https://doi.org/10.1016/B978-0-12-372560-8.X5000-1

Power, J. D., Mitra, A., Laumann, T. O., Snyder, A. Z., Schlaggar, B. L., & Petersen, S. E. (2014). Methods to detect, characterize, and remove motion artifact in resting state fMRI. *NeuroImage*, *84*, 320–341. https://doi.org/10.1016/J.NEUROIMAGE.2013.08.048

Satterthwaite, T. D., Elliott, M. A., Gerraty, R. T., Ruparel, K., Loughead, J., Calkins, M. E., Eickhoff, S. B., Hakonarson, H., Gur, R. C., Gur, R. E., & Wolf, D. H. (2013). An improved framework for confound regression and filtering for control of motion artifact in the preprocessing of resting-state functional connectivity data. *NeuroImage*, *64*(1), 240–256. https://doi.org/10.1016/J.NEUROIMAGE.2012.08.052

Tustison, N. J., Avants, B. B., Cook, P. A., Zheng, Y., Egan, A., Yushkevich, P. A., & Gee, J. C. (2010). N4ITK: Improved N3 bias correction. *IEEE Transactions on Medical Imaging*, *29*(6), 1310–1320. https://doi.org/10.1109/TMI.2010.2046908

Whitfield-Gabrieli, S., & Nieto-Castanon, A. (2012). Conn: a functional connectivity toolbox for correlated and anticorrelated brain networks. *Brain Connectivity*, *2*(3), 125–141. https://doi.org/10.1089/brain.2012.0073

Zhang, Y., Brady, M., & Smith, S. (2001). Segmentation of brain MR images through a hidden Markov random field model and the expectation-maximization algorithm. *IEEE Transactions on Medical Imaging*, *20*(1), 45–57. https://doi.org/10.1109/42.906424
